# Calibrated prediction intervals for polygenic scores across diverse contexts

**DOI:** 10.1101/2023.07.24.23293056

**Authors:** Kangcheng Hou, Ziqi Xu, Yi Ding, Arbel Harpak, Bogdan Pasaniuc

**Affiliations:** Bioinformatics Interdepartmental Program, University of California, Los Angeles, Los Angeles, CA, USA; Department of Computer Science, University of California, Los Angeles, Los Angeles, CA, USA; Department of Population Health, The University of Texas at Austin, Austin, TX, USA; Department of Integrative Biology, The University of Texas at Austin, Austin, TX, USA; Department of Pathology and Laboratory Medicine, David Geffen School of Medicine, University of California, Los Angeles, Los Angeles, CA, USA; Department of Computational Medicine, David Geffen School of Medicine, University of California, Los Angeles, Los Angeles, CA, USA; Institute for Precision Health, University of California, Los Angeles, Los Angeles

## Abstract

Polygenic scores (PGS) have emerged as the tool of choice for genomic prediction in a wide range of fields from agriculture to personalized medicine. We analyze data from two large biobanks in the US (All of Us) and the UK (UK Biobank) to find widespread variability in PGS performance across contexts. Many contexts, including age, sex, and income, impact PGS accuracies with similar magnitudes as genetic ancestry. PGSs trained in single versus multi-ancestry cohorts show similar context-specificity in their accuracies. We introduce trait prediction intervals that are allowed to vary across contexts as a principled approach to account for context-specific PGS accuracy in genomic prediction. We model the impact of all contexts in a joint framework to enable PGS-based trait predictions that are well-calibrated (contain the trait value with 90% probability in all contexts), whereas methods that ignore context are mis-calibrated. We show that prediction intervals need to be adjusted for all considered traits ranging from 10% for diastolic blood pressure to 80% for waist circumference. Adjustment of prediction intervals depends on the dataset; for example, prediction intervals for education years need to be adjusted by 90% in All of Us versus 8% in UK Biobank. Our results provide a path forward towards utilization of PGS as a prediction tool across all individuals regardless of their contexts while highlighting the importance of comprehensive profile of context information in study design and data collection.

## Introduction

Accurate prediction of complex diseases/traits integrating genetic and non-genetic factors is essential for a wide range of fields from agriculture to personalized genomic medicine. The genetic contribution to common traits is typically predicted using polygenic scores (PGS) that summarize the joint contribution of many genetic factors^1–4^. A critical barrier in PGS use is their *context-specific accuracy* – their performance (and/or bias) varies across genetic ancestry^5–9^, age, sex, socioeconomic status and other factors^10–12^. This prevents equitable use of PGS across individuals of all contexts^4, 5, 13^.

PGS use data from large-scale genome-wide association studies (GWAS) to estimate linear prediction models of traits based on genetic variants; these prediction models are then used for new data that often has different context characteristics from the GWAS training data (e.g., different distributions of genetic ancestry, social determinants of health, etc.)^1, 2, 14^. Even when testing data is similar to training data, genetic effects themselves can vary by contexts (e.g., due to genotype-environment interaction, across age^15^, sex^16^, genetic ancestry^17–20^) thus leading to differential PGS performance (as traditional PGS do not model such interactions). Furthermore, when genetic effects are unknown, allele frequency, linkage disequilibrium and differential tagging of true latent genetic factors can also lead to context-specific accuracy of PGS-based predictions^10, 15, 21^.

To account for PGS accuracy variability, we propose an approach to incorporate context-specificity into *trait prediction intervals* that are allowed to vary across contexts. Trait prediction intervals denote the range containing true trait values with pre-specified confidence (e.g., 90%). And they provide a natural approach to model variability in PGS accuracies – narrower prediction intervals correspond to contexts where PGS attains higher accuracy – that can then be used in applications of PGS-based trait predictions^10, 22, 23^. As an example, consider the case of two individuals with the same PGS-based predictions for low-density lipoprotein cholesterol (LDL) of 120 mg/dL. If the two individuals have different contexts (e.g., sex) that are known to impact PGS accuracy (e.g., *R*^2^=0.1 in men vs. 0.2 in women), their prediction intervals will also vary (e.g., 120 ± 40 mg/dL vs. 120 ± 10 mg/dL). In this example the second individual is more likely to meet a decision criterion of LDL>100 mg/dL for clinical intervention.

To achieve calibration across all contexts, we propose a statistical model (*CalPred*) that jointly models the effects of all contexts on PGS accuracy leveraging calibration data. The key assumption is that new target individuals for whom PGS-based predictions will be employed have similar contexts as the calibration data. This is motivated by precision health efforts that created EHR-linked biobanks of patients from the same medical system in which the PGS-based prediction will be applied in the future^24–27^; in this context the assumption is that the biobank is representative of future patients entering the same medical system.

First, we analyze data across two large-scale biobanks (UK Biobank^28^ and All of Us^29^) to find pervasive impact of context on PGS accuracy across a wide range of traits. All considered traits (N=72) have at least one context impacting their accuracies^10, 12^. Socio-economic contexts have similar magnitudes of impact on PGS accuracies as genetic ancestry; for example, PGS accuracy varies by up to ∼50% for individuals across the context of “education years” averaged across all considered traits in All of Us. Moreover, socio-economic contexts have greater impact on PGS accuracy in All of Us, a more diverse dataset, as compared to UK Biobank.

Second, we use simulations and real data analysis to find that CalPred provides well-calibrated prediction intervals across individuals of diverse contexts. For example, CalPred jointly models the impact of genetic ancestry, age and sex and other social determinants of health on LDL prediction to find that prediction intervals need adjustment by up to ∼40% across contexts to achieve calibration. The context-specificity of PGS prediction varies across traits, with largest adjustments observed for traits including waist circumference and average mean spherical equivalent (avMSE) where prediction intervals need adjustment by ∼100% for individuals in certain contexts; meanwhile certain traits such as diastolic blood pressure only need a modest adjustment by ∼20%. Notably, the context-specificity of the same trait also depends on the studied population; for example, prediction intervals for education years need adjustment by 90% in All of Us versus 8% in UK Biobank, reflecting the more diverse distribution of education years and other social determinants of health in All of Us. Overall, our approaches provide a path forward to modeling differential PGS accuracy by context in prediction of complex traits in humans.

## Results

### Overview

We incorporate context-specific accuracy in PGS-based predictions using prediction intervals that are allowed to vary across contexts to maintain calibration: the true phenotype is contained within the prediction interval at a pre-specified probability (e.g., 90%; Figure 1a). Naturally, as accuracy varies by context, the interval width needs to vary adaptively such that calibration is maintained (Figure 1b). For illustrative purpose we distinguish among three types of prediction intervals (Figure 2). First, standard errors of PGS weights can be used to estimate prediction intervals that do not vary across contexts and/or individuals; these types of intervals are calibrated only when target perfectly matches training which is hard to achieve in practice. Second, prediction intervals can be estimated empirically using a calibration dataset across all data ignoring context^1, 30–34^; these types of intervals are robust to mismatches between training and testing, but are mis-calibrated in particular contexts due to the variability of PGS accuracy. Third, prediction intervals that vary across contexts can be estimated using a calibration dataset by empirically quantifying the impact of each context on prediction accuracy; context-specific prediction intervals are adaptive and robust across contexts albeit at the expense of a more complex statistical model and larger calibration data that spans all contexts. Motivated by clinical implementation of PGS-based predictions in medical systems where EHR-linked biobanks already exist, here we focus on leveraging calibration data to estimate context-specific prediction intervals. In this scenario it is natural to use existing EHR-linked biobanks as approximation for future patients within the same medical system. For example, UCLA ATLAS biobank^24^ contains data of ∼150k patients within the UCLA Health system that can be used to calibrate PGS-based predictors for future visits of UCLA patients.

**Figure 1:**
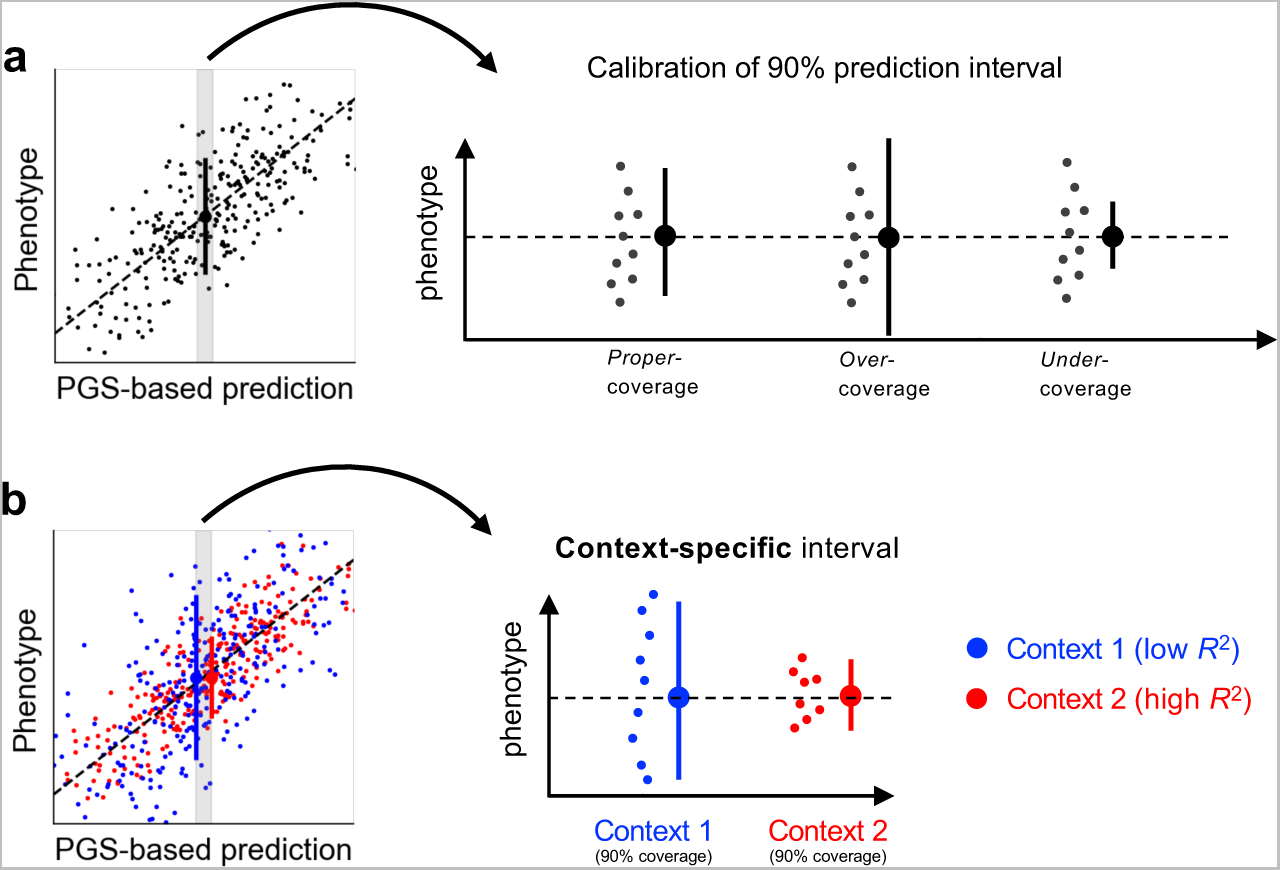
Calibrated and context-specific prediction intervals via CalPred. **(a)** Calibration of prediction intervals. We consider a subset of individuals with the same point prediction (shaded area in the left panel, dashed horizontal line in the right panel). Each dot denotes an individual’s phenotype value. Intervals with *proper-coverage* cover the true phenotype at pre-specified probability of 90%; intervals with *over-coverage* are incorrectly wide; intervals with *under-coverage* are incorrectly narrow. **(b)** Context-specific calibration of prediction intervals. We consider two subpopulations in different contexts (e.g., female and male). Context 1 (blue dots) has lower prediction accuracy and therefore wider variation around the mean, while context 2 (red dots) has higher prediction accuracy and therefore narrower variation around the mean. Context-specific intervals vary by context, providing intervals with proper coverage in each context.

**Figure 2:**
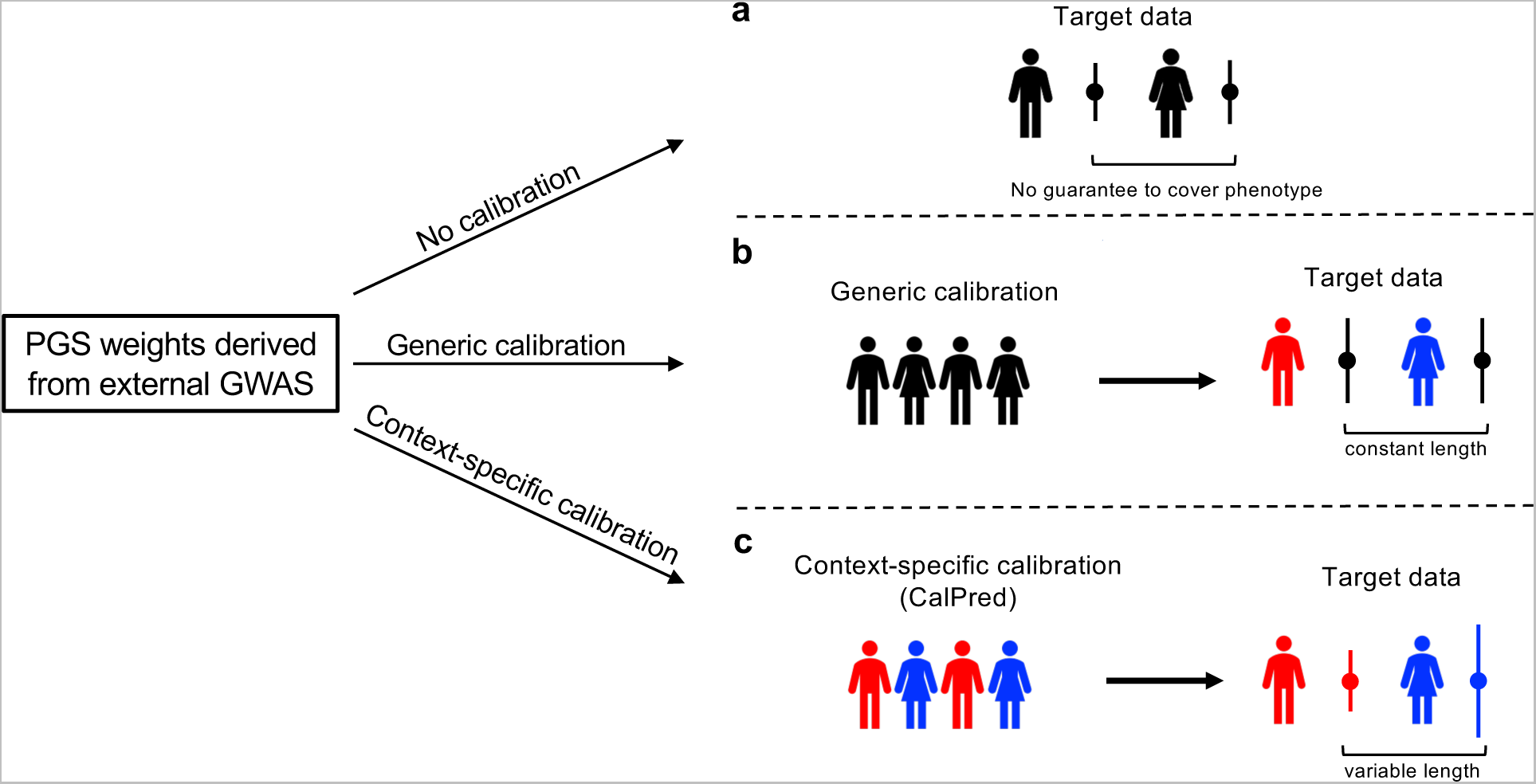
Different approaches for prediction intervals of PGS-based models. All approaches start with a set of predefined PGS weights derived from existing GWAS. **(a)** prediction intervals can be calculated using analytical formula without calibration data. However, these intervals are not guaranteed to be well-calibrated. **(b)** Generic calibration methods do not consider context information; they produce generic prediction intervals that are constant across individuals. **(c)** Context-specific calibration leverages a set of calibration data to estimate the impact of each context to trait prediction accuracy; the estimated impact can then be used to generate prediction intervals for any target individuals matching in distribution with calibration data.

Mathematically, we model context-specific prediction accuracy via the error term 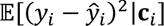 for phenotype 𝑦*_i_* and prediction mean (or point prediction) 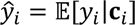 as a function of context 𝐜_!_for each individual *i* in the calibration dataset. We parametrize the impact of all contexts on prediction intervals in a joint model as 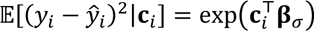 where 𝐜_i_denotes contexts including age, sex, socioeconomic factors and top principal components (denoting major axes of genetic ancestry; Methods). 𝛃_$_quantifies the unique impact of each context on variation of the prediction interval accounting for other contexts (Methods). This approach is a generalization of the context-free approach. Denoting prediction standard deviation (SD) as 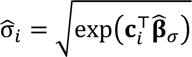, 90% prediction intervals can be derived as 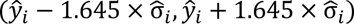.

### Widespread context-specific PGS accuracy in diverse populations

Although PGS accuracy has been shown to vary across selected traits and contexts^5, 10–12^, its pervasiveness remains unclear. We analyzed two large-scale biobanks in the UK and US (UK Biobank and All of Us) comprising >600K individuals spanning a wide range of contexts. We trained PGS for 72 traits in individuals previous annotated as “White British”^28^ (WB) from UK Biobank and evaluated these PGSs in independent testing data from UK Biobank and All of Us. We focused on 11 contexts that span genetic ancestry, sex, age, and socio-economic factors such as educational attainment (Methods). We used *relative* Δ*R*^2^ to quantify the impact of context to PGS accuracy defined as 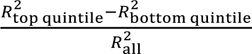, where 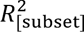 denotes *R*^2^ between PGS and residual phenotype computed in a given range of the context variable (top/bottom quintile for continuous contexts; binary subgroups for binary contexts). We found widespread context-specific PGS accuracies across all traits and contexts studied (Figure 3, S1 and S2, Table S1 and S2; Methods).

**Figure 3:**
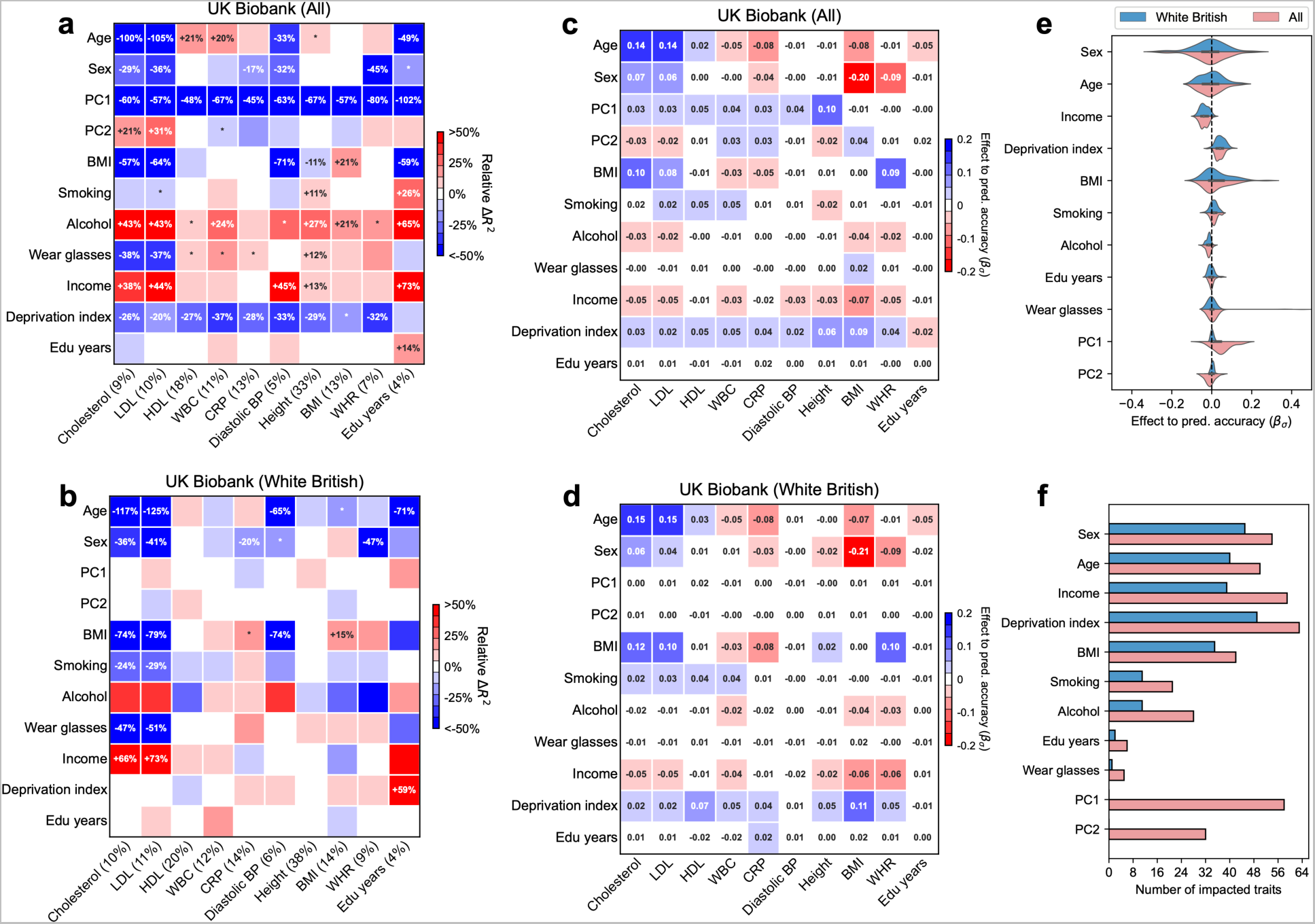
Widespread context-specific PGS prediction accuracy in UK Biobank. **(a-b)** Heatmaps for context-specific PGS accuracy for all and WB individuals. Each row denotes a context and each column denotes a trait; the squared correlation between PGS and residual phenotype (*R*^2^) is shown in parentheses. Heatmap color denotes the PGS-phenotype relative Δ*R*^2^ (defined as 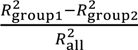 , where 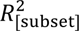 represents *R*^2^ computed in a given range of the context variable. For continuous contexts, relative Δ*R*^2^ denote differences of top quintile minus bottom quintile; for binary contexts (including sex, smoking, wear glasses, alcohol), relative Δ*R*^2^ denote differences of male minus female, smoking minus not smoking, wearing glasses minus not wearing glasses, drinking alcohol minus not drinking alcohol (these orders were arbitrarily chosen). Numerical values of relative *R*^2^ differences are displayed for PGS-context pairs with statistically significant differences (multiple testing correction for all 10×11 PGS-context pairs in this figure; *p* < 0.05 / (10×11)). ‘*’ are displayed for PGS-context pairs with nominally significant differences (multiple testing correction for 11 contexts; *p* < 0.05 / 11). **(c-d)** Heatmaps for effects to prediction accuracy in CalPred model (estimated 𝛽_+_ ). Colormaps were inversed to those of **(a-b**) to reflect that positive 𝛽_+_ corresponds to lower prediction accuracy and vice versa. **(e)** Distribution of estimated 𝛽_+_ in the CalPred model for each context across traits. **(f)** Number of significantly impacted traits by each context (*p* < 0.05 / (72×11)).

### Context-specific accuracy in UK Biobank

All 72 traits had at least one context impacting their accuracies in UK Biobank data; 264 (out of 792) PGS-context pairs had significant variable accuracy (*p* < 0.05 / (72 × 11); Methods). Overall, genetic ancestry had the most widespread impact on PGS accuracy: 70 of 72 traits had significant differences in PGS accuracy, with an average relative Δ*R*_2_ of −46% between top and bottom PC1 quintiles (Figure S3). Socioeconomic contexts also significantly impacted PGS accuracy; PGS accuracy significantly differed for 62 traits, with an average relative Δ*R*^2^ of −23% between top and bottom deprivation index quintiles. The direction of context’s impact depended on the trait being studied. For example, age significantly impacted 19 traits; rather than consistently increasing or decreasing accuracy, an older age led to increased accuracies for 13 traits (e.g., high-density lipoprotein cholesterol and white blood cell count in Figure 3; HDL and WBC) and to decreased accuracies for 6 traits (e.g., low-density lipoprotein cholesterol; LDL).

The widespread context-specificity retained even when testing data was matched to the training data by genetic ancestry (Figure 3). 22 (out of 72) PGSs had at least one context significantly impacting their prediction accuracies; 43 PGS-context pairs had significant variable accuracy (*p* < 0.05 / (72 × 11)). We replicated previously reported variable PGS accuracy in WB individuals for diastolic blood pressure, body mass index, education years across contexts of sex, age and deprivation index^10^. As an example, LDL was significantly impacted by six contexts in WB individuals, with age having the strongest impact (relative Δ*R*^2^ was more than 100% between top and bottom age quintiles).

Next, we studied the unique impact of each context on variable PGS accuracy within CalPred model that jointly accounts for all contexts (Methods, Figure 3cd). Context contribution to variable accuracy conditional on all other contexts was quantified with 𝛽_$_ , where larger absolute 𝛽_$_ indicated more substantial variation in accuracy along a context variable (Methods). In general, the effects of contexts to traits were largely independent. For example, both PC1 and deprivation index significantly impacted PGS accuracy for a range of traits in the joint model, indicating both had a unique contribution to variable PGS accuracy. We also found examples showing otherwise: the impact of “wear glasses” context on LDL accuracy can be explained by its correlation with age (Figure S4), while other contexts independently contributed to variable LDL accuracy. These results indicated the importance of jointly considering all measured contexts to correctly assess the unique contribution of each context. We found that contexts including sex, age, income, and deprivation index had comparable impact on accuracy as genetic ancestry (Figure 3ef). The distribution of estimated effects of 𝛽_$_suggested predominantly higher prediction accuracy for individuals with higher income and lower deprivation indices; this can be partly explained by different context distribution PGS training data: WB individuals had higher income and lower deprivation indices compared to the rest of the UK Biobank^35^ (Figure S5).

### Context-specific accuracy in All of Us

We next turned to All of Us, a diverse biobank across the US comprising more than 160K participants (Figure S3 and S6). Due to challenges in phenotype matching across biobanks, we restricted the analysis to 10 traits and 11 contexts matching the UK Biobank analyses (Methods). All traits had at least one context that impacted their accuracies (Figure 4, Table S3 and S4). 81 PGS-context pairs were significant when considering all individuals, and 49 PGS-context pairs were significant when restricting to individuals with self-reported race/ethnicity (SIRE) as “White” (“White SIRE”) (*p* < 0.05 / (12 × 11); Methods). Prediction of cholesterol and LDL were similarly impacted by a broad range of contexts. Prediction of education years was impacted by contexts including age, BMI, employment, income, both when considering all individuals and considering “White SIRE” sample, consistent with evidence that socioeconomic contexts influence PGS of socio-behavioral traits such as education^10, 36, 37^.

**Figure 4:**
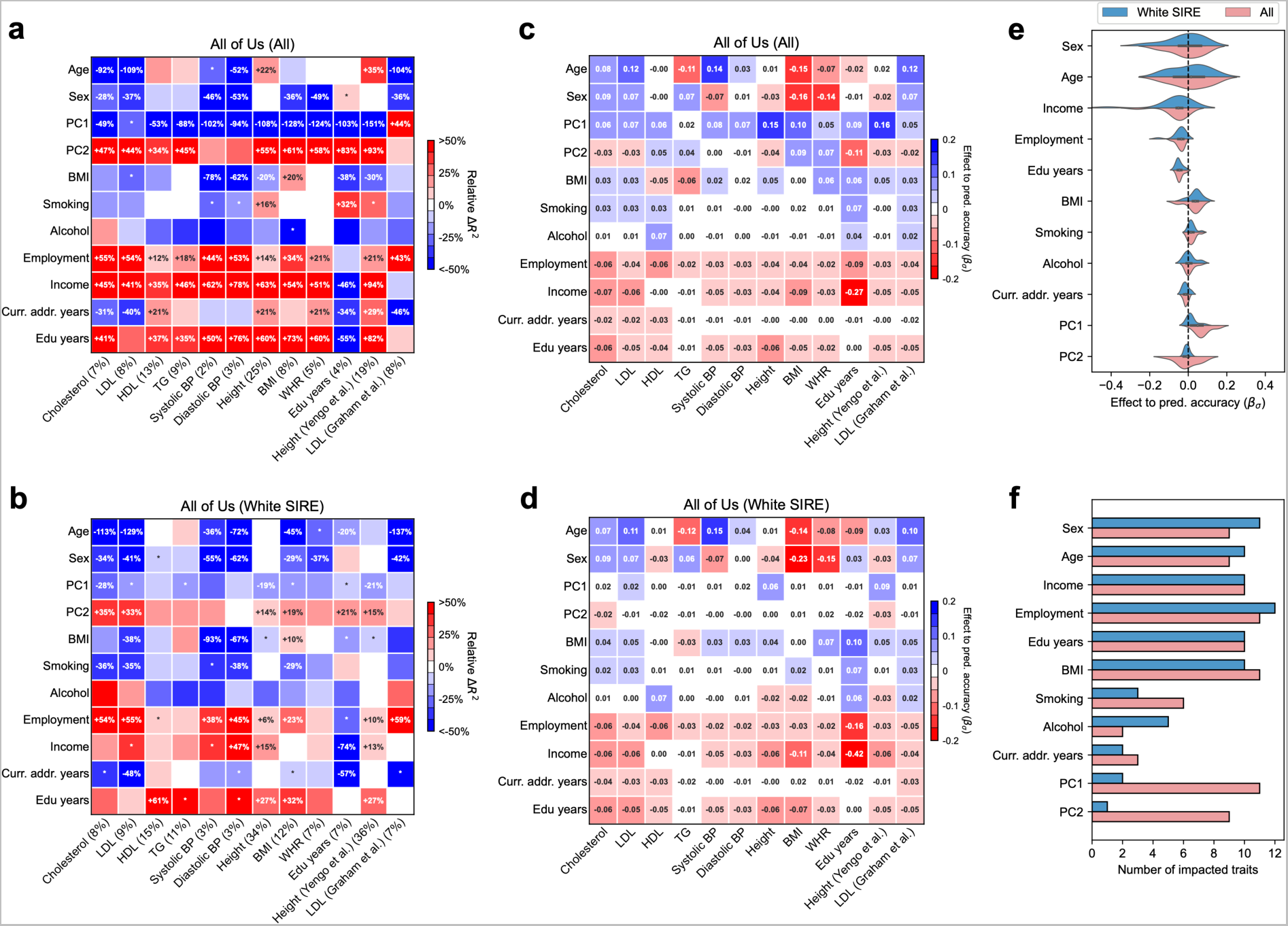
Widespread context-specific PGS prediction accuracy in All of Us. **(a-b)** Heatmaps for context-specific PGS accuracy for all and white SIRE individuals. Each row denotes a context and each column denotes a trait; overall *R*^2^ is shown in parentheses. Heatmap color denotes relative Δ*R*^2^: differences of top quintile minus bottom quintile for continuous contexts and difference of male minus female for binary context of sex. Numerical values of relative *R*^2^ differences are displayed for trait-context pairs with statistically significant differences (multiple testing correction for all 12×11 trait-context pairs in this figure; *p* < 0.05 / (12×11)). ‘*’ are displayed for trait-context pairs with nominally significant differences (multiple testing correction for 11 contexts; *p* < 0.05 / 11). **(c-d)** Heatmaps for estimated 𝛽_+_ in CalPred model. **(e)** Distribution of estimated 𝛽_+_in CalPred model for each context across traits. **(f)** Number of significantly impacted traits by each context (*p* < 0.05 / (12×11)).

Interestingly, socioeconomic contexts had greater impact on context-specificity in All of Us as compared to UK Biobank. For example, years of education context significantly impacted 9 out of 11 traits with average relative Δ*R*^2^=50%, as compared to 2 out of 71 traits with average relative Δ*R*^2^=0.2% in UK Biobank (averaging across traits other than education years itself). This may be explained by larger variation of education years in the US and/or education being more correlated with latent social determinants of health in the US as compared to the UK.

For completeness we also evaluated PGSs for height^38^ and LDL^39^ derived from multi-ancestry meta-analyses from PGS Catalog^40^ (Figure 4). We found that multi-ancestry PGSs did not alleviate widespread context-specific accuracy. Higher income, education years, better employment, or lower BMI predominately led to higher prediction accuracy across traits (Figure 4ef). We formally compared and determined an overall consistency for fitted 𝛽_$_ coefficients across populations and biobanks (Figure S7). We determined that variable *R*^2^ across contexts was not solely driven by differences of phenotype variance in context strata: context-specific *R*^2^ can result from differences in either phenotypic variance or PGS predictiveness, and the extent attributed to either component varied by each context-trait pair (Figure S8).

### CalPred yields calibrated context-specific prediction in simulations

Having shown that context-specificity of PGS accuracy is pervasive across traits and biobanks, we next turned to CalPred, an approach to estimate context-specific prediction intervals accounting for context-and trait-specific variable accuracy (Methods). We first evaluated CalPred in simulations where prediction accuracy varies across contexts similar to real data^5, 6, 10^ (Figure 5; Methods). We assessed calibration of prediction intervals at both the overall level and within each context subgroup (Methods). First, we showed that generic prediction intervals without context-specific adjustment had severe over-/under-coverage when evaluated within each context subgroup stratified by PC1, age, or sex. As expected, biases of coverage tracked closely with accuracy across contexts (Figure 5). Second, we showed that CalPred context-specific prediction intervals that were allowed to vary with each individual’s context were calibrated across contexts (Figure 5). This was due to the incorporation of context-specific prediction accuracy in the interval estimation. CalPred performance depended on calibration sample size with *N*_cal_>500 for accurate model fitting (Figure S9). Next, we investigated the impact of unmeasured context and found that CalPred was not calibrated across subgroups of individuals defined by the unmeasured context. In simulations where we included excessive contexts that did not impact prediction accuracy, coverages of prediction intervals were associated with larger standard errors, highlighting the importance of selecting an appropriate set of contexts in calibration (Figure S9). We also determined that parameter estimations of 𝛃_𝜎_ were accurate when the model was correctly specified and remained robust in model mis-specification scenarios (Figure S10). Overall, simulation results demonstrated that CalPred is able to produce well-calibrated and context-specific prediction intervals when contexts are measured and present in the data, and highlighted the importance of comprehensive profiling of relevant context information.

**Figure 5:**
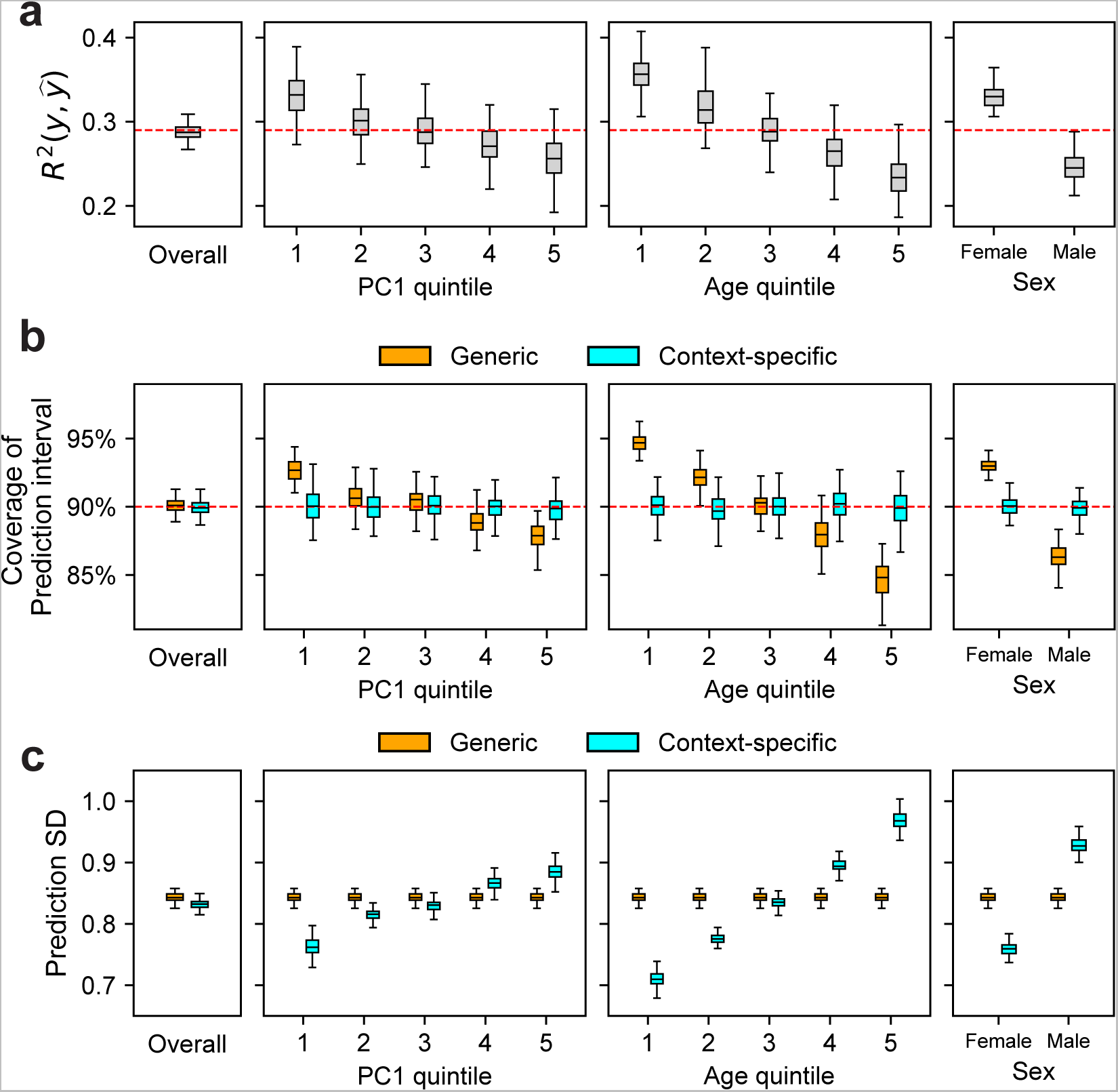
Simulation studies of CalPred. Simulations were performed to reflect scenarios where individuals have variable prediction accuracy by genetic PC1, age, and sex. For each simulation, we first trained a calibration model using a random set of 5,000 training individuals and then evaluated resulting prediction intervals on 5,000 target individuals (Methods). **(a)** Prediction *R*^2^ between 𝑦 and 𝑦% in simulated data both at the overall level, and in each context subgroup. **(b)** Coverage of generic vs. context-specific 90% prediction intervals evaluated in each context subgroup. Generic intervals were obtained by applying CalPred without context information; context-specific intervals were obtained by applying CalPred together with context information. **(c)** Average length of generic vs. context-specific prediction standard deviation (SD) in each context. Each box plot contains *R^2^*/coverage/average length evaluated across 100 simulations (100 points for each box plot), the center corresponds to the median; the box represents the first and third quartiles of the points; the whiskers represent the minimum and maximum points located within 1.5× interquartile range from the first and third quartiles, respectively.

### CalPred yields calibrated context-specific predictions in real data

Next we applied CalPred to produce context-specific prediction intervals for a wide range of traits across UK Biobank and All of Us. We start by showcasing LDL, an important risk factor of cardiovascular disease^39^. Calibration by context is particularly important because accuracy of predicting LDL was impacted by many contexts, with largest impact from age (Figure 3 and 4). We modeled the prediction mean using PGS together with age, sex, and genetic ancestry, and modeled context-specific prediction intervals using the set of contexts investigated in Figure 3 and 4 (Methods). Accuracy of LDL prediction decreased with age (*R*^2^=17% in youngest quintile vs. *R*^2^=11% in oldest quintile; Figure 6a). Generic prediction intervals were mis-calibrated with coverage of 93% and 86% for youngest and oldest quintiles instead of the nominal level of 90%. In contrast, context-specific prediction intervals had the expected 90% coverage across all considered contexts. This resulted from varying prediction interval length by context, with a wider interval compensating for lower prediction accuracy. For example, as the model estimated a positive impact of age to prediction uncertainty (𝛽_$_=0.15; *p*<10^-30^), individuals in youngest/oldest age quintiles had average prediction standard deviation (SD) of 27.9 vs. 34.5 mg/dL (24% difference; Figure S11; Methods). These findings were replicated in All of Us and in other traits (Figure S12 and S13), where *R*^2^ varied across contexts and context-specific prediction intervals achieved well-calibration.

**Figure 6:**
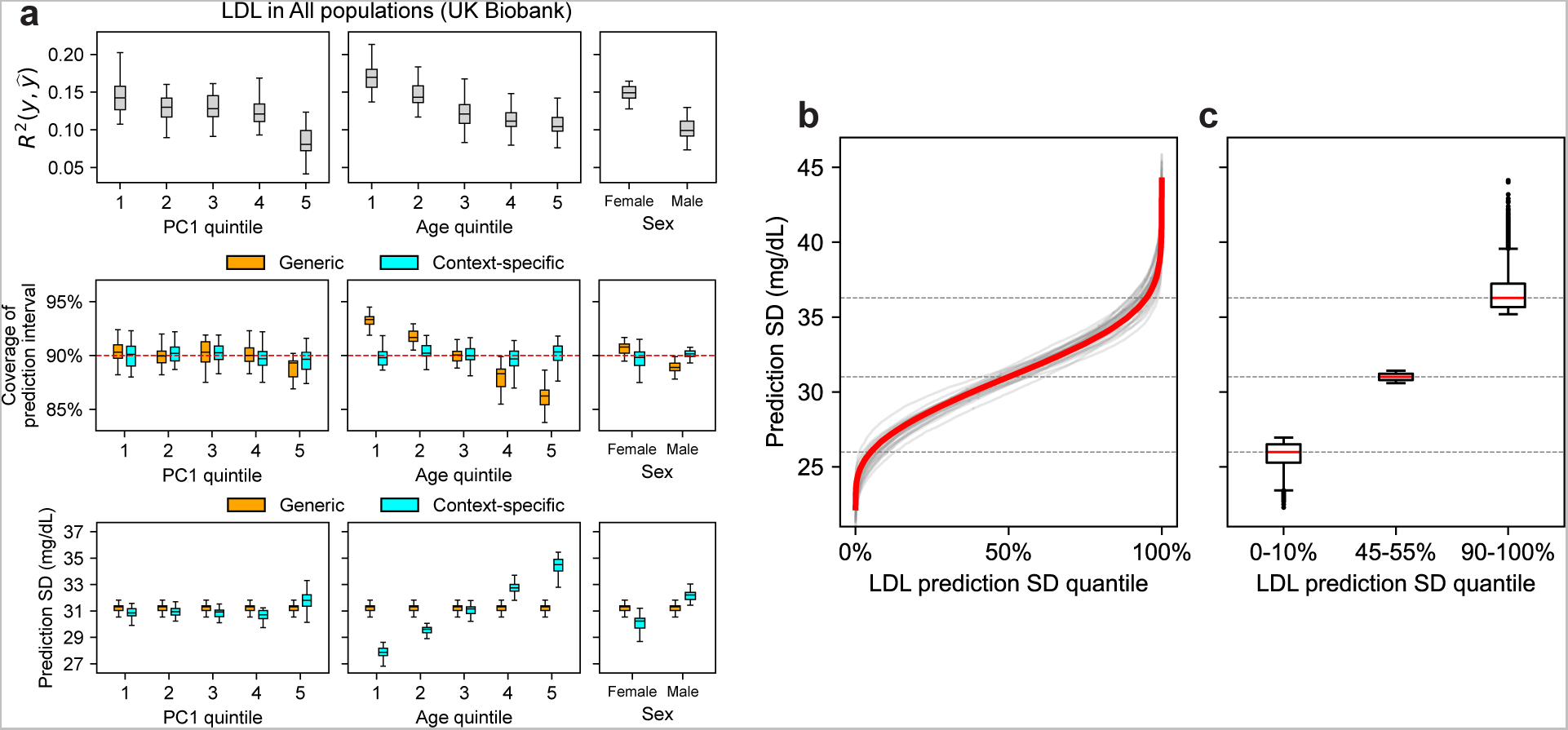
CalPred PGS calibration of LDL in UK Biobank. (a) Top panel: Prediction *R^2^*between phenotype and point predictions (incorporating PGS and other covariates) both at the overall level, and in each subgroup of individuals stratified by context. **Middle panel:** Coverage of generic vs. context-specific 90% prediction intervals evaluated in each context subgroup. Generic intervals were obtained by applying CalPred without context information; context-specific intervals were obtained by applying CalPred together with context information. **Bottom panel:** Average length of generic vs. context-specific 90% prediction intervals in each context. Each box plot contains *R^2^*/coverage/average length across 30 random samples with each sample of 5,000 training individuals and 5,000 target individuals (30 points for each box plot) **(b)** Ordered LDL prediction SD in unit of mg/dL. Gray lines denote prediction SD obtained with random sample of 5,000 training and applied to 5,000 target individuals. Red line denote prediction SD obtained from all individuals. **(c)** Box plots of results in (b) from individuals of LDL prediction SD quantile of 0-10%, 45-55%, 90-100%; the center corresponds to the median; the box represents the first and third quartiles of the points; the whiskers represent the minimum and maximum points located within 1.5× interquartile ranges from the first and third quartiles, respectively.

Next, we sought to examine the joint contribution of all considered contexts to variable prediction SD (instead of separately considering age, PC1 or sex; Figure 6b). Context-specific accuracy was more pronounced by ranking individuals by prediction SD accounting for impact of all contexts (prediction SD ranged approximately from 20 mg/dL to 45mg/dL; Figure 6b): we detected a 39% difference comparing individuals in bottom and top deciles of prediction SD (26.0 mg/dL vs. 36.3 mg/dL; Figure 6c; Figure S14 and S15). This implied that individuals in top prediction SD decile (characterized by contexts of male, increased PC1 and age; see LDL column in Figure 4c) needs to have their prediction interval widths increased by 39% compared to those in bottom decile.

Extending analysis accounting for all contexts to all traits in UK Biobank and All of Us, we determined a widespread large variation of context-specific prediction intervals across traits (Figure 7). Average differences between top and bottom prediction SD deciles across traits were 31% and 43%, respectively for UK Biobank and All of Us. The trait with the highest prediction SD difference was the average mean spherical equivalent (avMSE), a measure of refractive error, that was impacted the most by “wear glasses” context. Individuals who wore glasses had a much higher PGS-phenotype *R*^2^ (9.6%) than those who did not (2.2%), likely due to the reduced variation in avMSE phenotypes among individuals who did not wear glasses. Comparing across the two datasets, BMI, LDL, and cholesterol were more heavily influenced by context than average, while diastolic blood pressure and HDL were less impacted, suggesting trait-specific susceptibility to context-specific accuracy. Notably, there were also cases where context-specificity of the same trait was drastically different across datasets. For example, prediction SD differences for predicting education years was 90% in All of Us versus 8% in UK Biobank. This disparity likely reflected the more diverse distribution of education years and other social determinants of health in the US population sampled in All of Us, in line with results in Figures 2 and 3. Such differences between datasets also highlight that context-specificity can be population-specific and the need to consider unique characteristics of different populations in calibration. Taken together, our findings emphasize the importance of incorporating context information into PGS-based models when applied in diverse populations.

**Figure 7.**
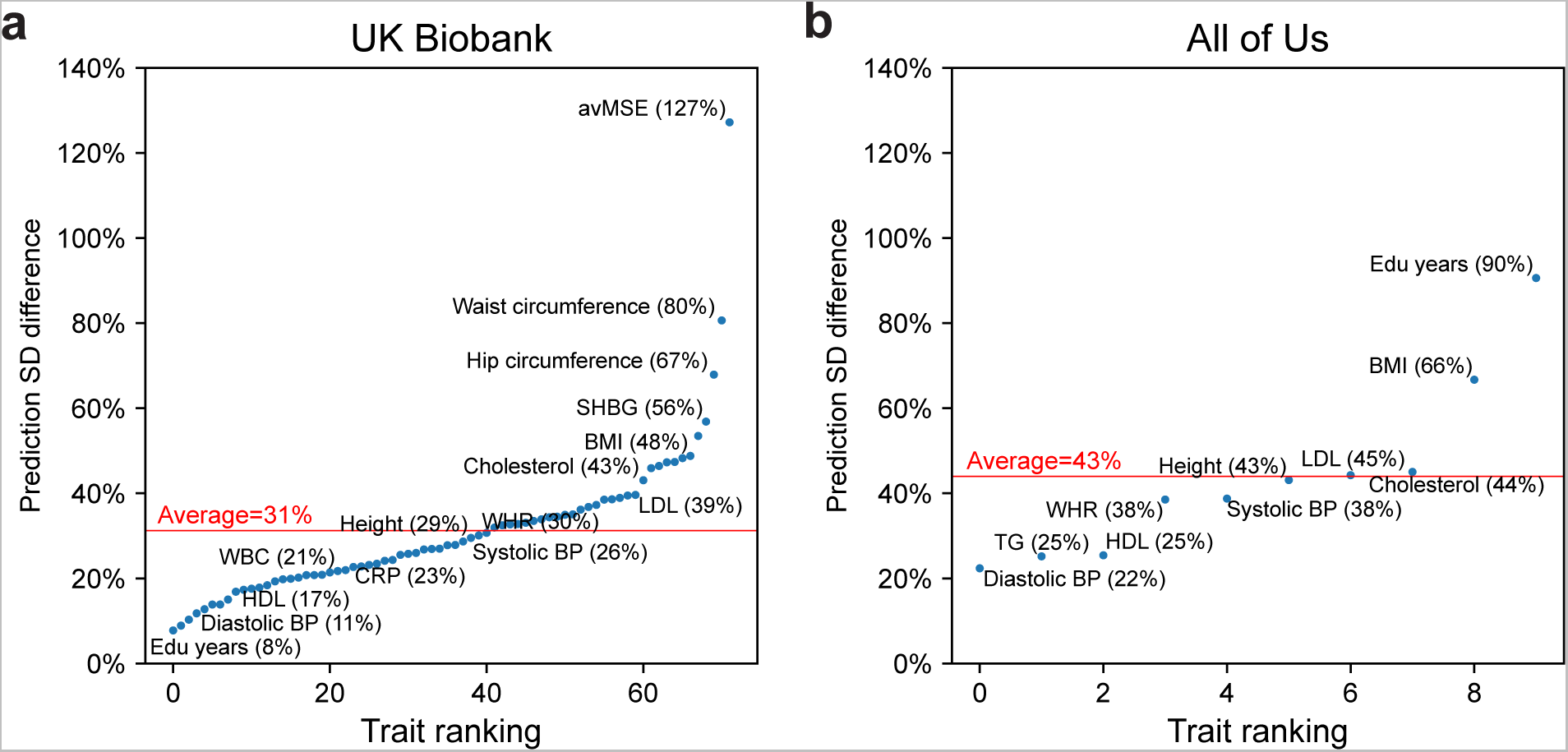
Variation of prediction standard deviation (SD) accounting for all contexts. Relative difference of prediction SD between top and bottom prediction SD deciles (90-100% vs. 0-10%) for all traits in UK Biobank **(a)** and All of Us **(b)**. Traits are ranked by prediction SD. The difference is calculated with the median prediction SD within decile of individuals with highest prediction SD 𝑆_𝑑1_ and decile of individuals with lowest prediction SD 𝑆_𝑑10._ Using 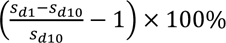 .

## Discussion

Our work adds to the literature of PGS-based prediction as follows. First, we show that context-specific accuracy of PGS is highly pervasive across traits and biobanks with socioeconomic contexts often having larger impact than genetic ancestry^5, 10, 12, 23, 41^. Second, we introduce CalPred to estimate context-specific prediction intervals that maintain calibration for all individuals across contexts. Third, we show using real and simulated data how differential prediction intervals can be used to incorporate uncertainty in predictions. Although we focused primarily on PGS-based prediction, our approaches are general and can incorporate any other factors. Fourth, we focused on trait prediction as the main output of our approach motivated by genomic medicine applications. As PGSs are increasingly applied to diverse populations, we find it imperative to incorporate the context-specific accuracy into PGS downstream analyses to avoid bias against certain contexts due to differential prediction accuracy, especially for contexts that are correlated with socioeconomic status. CalPred provides a principled framework to quantify generalizability/portability of a given PGS and represent individualized context-specific accuracy to be leveraged in downstream analyses. The prediction intervals can be interpreted as a personalized reference range accounting for each individual’s contexts (including age, sex, and genetic variation via PGS). Such personalized reference range may prove useful in identifying individuals with outlier lab values in a personalized and equitable fashion to prevent under-/over-diagonsis^42^.

The observation that distribution of PGSs differs across genetic ancestry continuum^41^ motivates methods that regress out effects of variables representing genetic ancestry from PGS distribution to facilitate comparison across individuals locating at different positions in genetic ancestry continuum^43, 44^. However, such approaches may unintentionally remove true biological differences of PGS distribution across genetic ancestry continuum (e.g., African Americans have reduced neutrophil count that can be explained by the large effect of a single Duffy-null SNP^45^) as they do not consider phenotype value distribution in calibration procedure; in addition, these approaches cannot represent different standard errors in PGS predictions of individuals across genetic ancestry continuum. Our method leverages a set of calibration data to properly adjust point predictions across contexts according to true phenotype distribution. Compared to other existing calibration methods^34^, our approach provides a framework to incorporate context information.

We note several limitations and provide future directions of our work. First, we focused on modeling and analyzing quantitative traits in this work. Context-specific accuracies can be further incorporated in modeling case-control status and absolute risk of diseases, perhaps by modeling the underlying disease liability using methods proposed in this study. Second, we made several modeling assumptions, including the linear relationship between error terms and contexts, as well as quantile normalization procedure to phenotype values to fit in normal assumption of CalPred model. Future work may leverage models with fewer assumptions and calibration dataset with larger sample size to enable more flexible modeling. Third, CalPred requires calibration data that matches in distribution with the target data, including both the distribution of contexts and their effects to phenotypes (in terms of both prediction mean and variance). Otherwise, there may be bias in target samples that are underrepresented in the calibration data. The magnitude of bias due to mismatch between calibration and target data in realistic scenarios needs to be empirically examined in future work. As shown in our simulation studies, missing contexts will also limit proper calibration of PGS along such contexts; this observation advocates standardized and comprehensive profiling of contexts across biobanks to better quantify the role of contexts to PGS accuracy, especially for those related to social-economic status, to prevent further exacerbation of health disparity. Relatedly, these results indicate that GWAS data collecting process not only needs to prioritize diversity in genetic ancestry, but also promote diversity across social-economic contexts, because PGS may be estimated with different precision in different social-economic contexts. Fourth, CalPred prediction intervals will benefit from improved modeling of the prediction mean; this may be achieved by more fine-grained modeling of prediction factors to capture more phenotype variation (Supplementary Note). For example, sex-specific SNP-level effects can be estimated from individual-level GWAS data^16^ and CalPred coupled with sex-specific PGS is likely to produce more precise, and shorter, prediction intervals.

## Methods

### Constructing calibrated and context-specific prediction intervals

We first provide an overview of CalPred framework. CalPred takes as input from pre-trained PGS weights, genotype, phenotype and contexts to train a calibration model to generate calibrated and context-specific prediction intervals for target individuals. We consider a calibration dataset with *N*_cal_ individuals. For each individual i=1, …, *N*_cal_, we have measured genotype vector 𝐠_𝑖_ ∈ {0,1,2}^𝑀^ with M SNPs, and phenotype 𝑦_𝑖_. With pre-trained PGS weights for a given trait 𝛃_𝑔_ ∈ ℝ^𝑀^, we calculate the PGS for everyone in the calibration data with 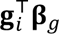. Each individual’s PGS, together with other contexts, including age, sex, genetic ancestry and other socioeconomic factors, compose each individual *i’*s contexts 𝐜_𝑖_ (all ‘1’ intercepts are also included). Phenotypes are then modeled as

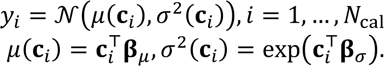

There are two main components in the model

- 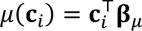 models the baseline prediction mean. This term is commonly used to predict phenotypes using PGS together with other contexts.
- 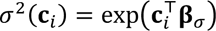 models the context-specific variance of 𝑦 around prediction mean. Differential prediction accuracy across contexts can lead to variable variance around prediction mean across contexts. The use of exp (⋅) is to ensure that the variance term >= 0.

Model parameters 𝛃_𝜇_, 𝛃_𝜎_ can be estimated leveraging a set of calibration data using restricted maximum likelihood for linear model with heteroskedasticity^46^ implemented in `statmod` R package^47^. Then individual-level predictive distribution 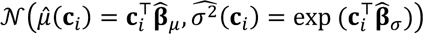 can be generated for any target individual 𝐜_𝑖_ using the fitted 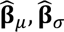. The corresponding 𝛼 -level prediction interval (e.g., 𝛼=90% for 90% prediction interval) is 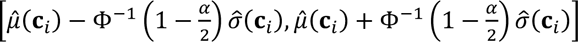, where Φ^−1^ is the inverse cumulative distribution function of a standard normal distribution (e.g., 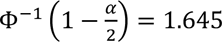 for 90% prediction interval). Since we fit a simple linear model, the extent of parameter overfitting is minimal with moderate sample size for calibration data (e.g., *N*_cal_>1,000 as validated in our simulation studies).

### Quantile normalization for non-normal phenotype distribution

In the above, we have assumed that prediction intervals can be properly modeled as a Gaussian distribution, which may not be always valid for every phenotype. To reduce the impact of this assumption to real data analysis, we apply a transformation function 𝑄(⋅) to 𝑦 with ranked based inverse normal transformation such that 𝑄(𝑦) follow a normal distribution; 𝑄(𝑦) can then be modeled using the methods described above. Fitted prediction intervals can then be transformed back into the original 𝑦 space using 𝑄^−1^(𝑦).

### Quantifying context-specific *R*^2^ of PGS

We quantify context-specific prediction accuracy (*R*^2^) of PGS, that is, to what extent PGS have variable prediction accuracy across contexts (including age, sex, genetic ancestry, proxies for lifestyle, socioeconomic contexts that can influence traits^48^). Accurate quantification of contexts contributing to variable prediction accuracy is important in constructing calibration model. In detail, for each pair of context and trait in a population, we calculated the prediction accuracy *R*^2^ between PGS 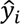 and covariate-regressed phenotypes 𝑦_𝑖_ (phenotypes for each trait were regressed out of age, sex, age*sex and top 10 PCs; this adjustment is to better separate the contribution of PGS) across each subgroup of individuals defined by contexts. We summarized results using relative differences of *R*^2^ across context groups to baseline *R*^2^ calculated across all evaluated individuals (relative differences between two classes for binary contexts; differences between top and bottom quintiles for continuous contexts). We calculated the Spearman’s *R*^2^ between point predictions and covariate-regressed phenotypes 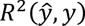 within each context subgroup. We also calculated the baseline Spearman’s *R*^2^ denoted as 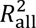 across all individuals regardless of contexts. We summarized the results for each pair of trait and context using the “relative Δ*R*^2^” defined as 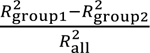. We assessed statistical significance of Δ𝑅^2^ across context subgroups by testing the null hypothesis 𝐻_0_: Δ𝑅^2^ = 0 using 1,000 bootstrap samples of Δ𝑅^2^ (in each bootstrap sample, the whole dataset was resampled with replacement and Δ*R*^2^ were then re-evaluated). Statistical significance was assessed using two-sided *p*-values comparing the observed Δ*R*^2^ to the bootstrap samples of Δ*R*^2^.

#### Relationship between CalPred model and *R*^2^

Population-level metrics such as *R*^2^ can be derived from this model as a function of 𝛃_𝜎_ and distribution of 𝐜_𝑖!_ Suppose 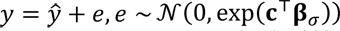 , where 𝑦, 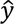 , 𝑒 denote the phenotypes, point predictions and residual noises, respectively. We have

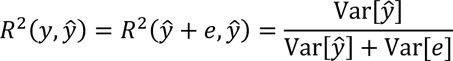

Holding 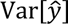 as fixed, 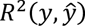 is a function of Var[𝑒], which is determined by the distribution of 𝐜 and values of 𝛃_𝜎_ . This indicates a correspondence between 𝛃_𝜎_ and 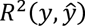]. Therefore, estimated 𝛃_𝜎_ can also be used as a metric to quantify context-specific accuracy (as used in Figure 3-4). While relative Δ*R*^2^ is easier to interpret, it assesses the marginal contribution of each context separately and require binning for continuous contexts. Meanwhile, 𝛽_𝜎_ in CalPred model jointly account for all contexts in parametric regression, and therefore can quantify the unique distribution of each context to variable accuracy.

On the other hand, even with constant prediction interval length (constant Var[𝑒]), variable 𝑅^2^ can still result from variable 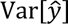 across context subgroups. While CalPred focus on modeling Var[𝑒] as a function of contexts to represent variable *R*^2^, 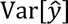 can also change as a function of contexts in certain scenarios. For example, 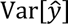 can vary with contexts if 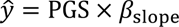 and the slope 𝛽 varies as a function of context. For example, ref.^16^ has reported 𝛽_slope_ can be different across contexts. Such variable slope term can be handled by modeling variable slope terms in prediction mean 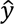 (Supplementary Note).

### Real data analysis

We analyzed a diverse set of contexts and traits in UK Biobank and All of Us (1) to quantify the extent of context-specific prediction accuracy and (2) to evaluate context-specific prediction intervals via CalPred.

#### Training polygenic score weights

Polygenic scores were trained on 370K individuals in UK Biobank that were assigned to “white British” cluster and 1.1M HapMap3 SNPs. For each trait, we performed GWAS using `plink2 --glm` with age, sex and top 16 PCs as covariates. Then we estimated PGS weights using `snp_ldpred2_auto` in LDpred2^49^ with input of GWAS summary statistics and in-sample LD matrix. These estimated PGS weights were then applied to target individuals in both UK Biobank and All of Us to obtain individual-level PGS. To train polygenic score weights to be used for individuals from All of Us, we overlapped 1.2M SNPs in All of Us quality-controlled microarray data to 12M SNPs in UK Biobank imputed data to obtain a set of 0.8M SNPs present in both datasets. Then we trained and applied polygenic scoring weights using these shared SNPs in UK Biobank to All of Us individuals. This procedure helps improve accuracy of the polygenic score in All of Us by ensuring all SNPs that have non-zero weights to present in the data.

#### UK Biobank dataset

We analyzed 490K genotyped individuals (including both training and target individuals). We used 1.1M HapMap3^50^ SNPs in all analyses. All UK Biobank individuals are clustered into sub-continental ancestry clusters based on top 16 pre-computed PCs (data-field 22009 in ref.^28^ as in ref.^6^). This procedure assigned 410K individuals into “white British” cluster. A random subset of 370K “white British” individuals to perform GWAS and estimate PGS weights (see above); we trained PGS weights starting with individual-level data to avoid overlap of sample between training and target data. For evaluation, we used the rest of 120K individuals with genotypes, phenotypes and contexts (including individuals from both ∼40K “White British” individuals and ∼80K other individuals). We focused on analyzing 72 traits with *R*^2^>0.05 in 40K WB target individuals and/or biological importance). We followed https://github.com/privefl/UKBB-PGS/blob/main/code/prepare-pheno-fields.R and ref.^6^ to perform basic preprocessing for trait values (e.g., log-transformation and clipping of extreme values). For each trait, we quantile normalized phenotype values; when performing calibration, phenotype quantiles were calculated based on calibration data and were then used to normalize target data. We analyzed 11 contexts representing a broad set of socioeconomic and genetic ancestry contexts, including binary contexts (sex, ever smoked, wear glasses, drinking alcohol) and continuous contexts (top two PCs, age, BMI, income, deprivation index, and education years). We note that income and education years have been processed into 5 quintiles in the original data of UK Biobank.

#### All of Us dataset

We analyzed 165K genotyped individuals with diverse genetic ancestry contexts (microarray data in release v6). We retained 1.2M SNPs from microarray data after basic quality control using plink2 with `plink2 --geno 0.05 --chr 1-22 --max-alleles 2 - -rm-dup exclude-all --maf 0.001`. We used microarray data because it contains more individuals and can be analyzed with low computational cost. All individuals with microarray data were used in the evaluation. We analyzed 10 heritable traits, including height, BMI, WHR, diastolic blood pressure, systolic blood pressure, education years, LDL, cholesterol, HDL, triglyceride; they are straightforward to phenotype and have large sample sizes. Physical measurement phenotypes were extracted from Participant Provided Information. Lipid phenotypes (including LDL, HDL, TC, TG) were extracted following https://github.com/all-of-us/ukb-cross-analysis-demo-project/tree/main/aou_workbench_siloed_analyses, including procedures of extracting most recent measurements per person, and correcting for statin usage. For each trait, we quantile normalized phenotype values; when performing calibration, phenotype quantiles were calculated based on calibration data and were then used to normalize target data. We included age, sex, age*sex, and top 10 in-sample principal components as covariates in the model. We also quantile normalized each covariate and used the average of each covariate to impute missing values in covariates. We analyzed 11 contexts, including binary contexts (sex) and continuous contexts (top two PCs, age, BMI, smoking, alcohol, employment, education, income, number of years living in current address).

#### Population descriptor usage

We explain our usage choices of population descriptor, including the use of top two PCs to capture genetic ancestry/similarity and the use of “white British” in analyses of UK Biobank and “white SIRE” in analyses of All of Us. We use the top two PCs computed across all individuals in UK Biobank or in All of Us, respectively, to capture the continuous genetic ancestry variation in each dataset. While these two PCs provide major axes of genetic variation (Figure S3), we acknowledge that top two PCs alone are not sufficient to fully capture all variation in the entire population. The discretized PC1 and PC2 subgroups used in Figure 2-6 is to enable calculation of population-level statistics such as 𝑅^2^ while we acknowledge that the underlying genetic variation is continuous. In UK Biobank, we intended to analyze a set of individuals with relatively similar genetic ancestry to perform GWAS and derive PGS. We used a set of individuals previously annotated with “white British” that were identified using a combination of self-reported ethnic background and genetic information having very similar ancestral backgrounds based on results of the PCA^28^. In All of Us, we selected a set of individuals with self-reported race/ethnicity (SIRE) being “white”, to study how PGS have different accuracy across environmental contexts in such a sample defined by SIRE. Noting that SIRE is not equivalent to genetic ancestry, the contrast of results from UK Biobank and All of Us helps understand how the genetic, nongenetic factors impact PGS accuracy in a group of individuals defined by SIRE or genetic ancestry.

#### Evaluating context-specific prediction intervals

Recall that the prediction mean and standard deviation are 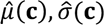 for a target individual with contexts 𝐜. We evaluate the prediction intervals with regard to phenotypes 𝑦 using metrics of

- Prediction accuracy: 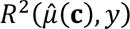.
- coverage of prediction intervals: evaluating 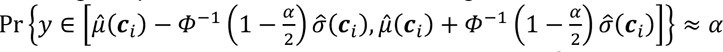, i.e., whether prediction intervals cover the true phenotypes with pre-specified probability of 𝛼.

Both metrics are evaluated both at the overall level for all individuals, and for each subgroup of individuals defined by contexts.

We generated and evaluated context-specific intervals in both UK Biobank and All of Us. For both datasets, we fit a model to simultaneously model the mean and variance where the mean term includes PGS, age, sex, age*sex, top 10 PCs so that this matches the baseline model that are commonly fitted, and the variance term includes age, sex, top 2 PCs, and other contexts of interest for each dataset (as shown in Figure 3-4 for UK Biobank and All of Us). For each trait, we performed the evaluation by repeatedly randomly sampling 5,000 individuals as calibration data to perform the calibration, and 5,000 individuals as target data to perform the evaluation (as described in “Constructing calibrated and context-specific intervals”).

### Simulations assessing coverage of context-specific prediction intervals

We simulated PGS point predictions 𝑦& and phenotype values 𝑦 to simulate traits with variable prediction accuracy across genetic ancestry continuum, age, and sex. We started with real contexts from 76K UK Biobank individuals not used for PGS training (see section “Real data analyses”). We used 3 contexts (PC1, age, and sex) in simulations. We quantile normalized each context so they had mean 0 and variance 1. Such simulations preserved the correlation between contexts. Given these processed contexts, we simulated point predictions 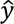 using a normal distribution 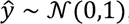 , and we simulated phenotypes 𝑦 with:

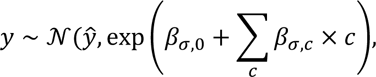

where 𝛽_𝜎,0_ denoted the baseline variance of 𝑦, and 𝛽_𝜎,𝑐_ was the effect of context 𝑐 to the variance of 𝑦. “Σ_𝑐_” enumerated over PC1, age, sex. This procedure simulated different variance of 𝑦 around 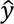 for individuals with different contexts, as observed in real data.

In details, we first selected 𝛽_𝜎,0_ such that 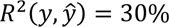 for individuals with average contexts (such that ∑_𝑐_ 𝛽_𝜎,𝑐_ × 𝑐 = 0 ). We simulated data with variable variances: we set 𝛽_𝜎,age_ = 0.25, 𝛽_𝜎,sex_ = 0.2, 𝛽_𝜎,PC1_ = 0.15. These parameters were manually chosen to roughly reflect the observed variable *R*^2^ in real data. In each simulation, we randomly sampled *N*_cal_=100, 500, 2500, 5000 individuals used for estimating the calibration model and *N*_test_ = 5000 individuals for evaluating the predictions from the set of 76K individuals. New point predictions and phenotypes 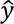, 𝑦 were simulated in each simulation. Then we quantified the prediction accuracy and coverage of prediction intervals in these simulations.

## Data availability

UK Biobank individual-level genotype and phenotype data are available through application at http://www.ukbiobank.ac.uk. AoU individual-level genotype and phenotype are available through application at https://www.researchallofus.org.

## Code availability

Software implementing CalPred and code for replicating analyses: https://github.com/kangchenghou/CalPred.

## Acknowledgements

We thank Molly Przeworski for helpful suggestions. This research was funded in part by the National Institutes of Health under awards R01HG009120, R01MH115676, and U01HG011715. This research was conducted using the UK Biobank Resource under applications 33127. We thank the participants of UK Biobank for making this work possible. The *All of Us* Research Program is supported by the National Institutes of Health, Office of the Director: Regional Medical Centers: 1 OT2 OD026549; 1 OT2 OD026554; 1 OT2 OD026557; 1 OT2 OD026556; 1 OT2 OD026550; 1 OT2 OD 026552; 1 OT2 OD026553; 1 OT2 OD026548; 1 OT2 OD026551; 1 OT2 OD026555; IAA #: AOD 16037; Federally Qualified Health Centers: HHSN 263201600085U; Data and Research Center: 5 U2C OD023196; Biobank: 1 U24 OD023121; The Participant Center: U24 OD023176; Participant Technology Systems Center: 1 U24 OD023163; Communications and Engagement: 3 OT2 OD023205; 3 OT2 OD023206; and Community Partners: 1 OT2 OD025277; 3 OT2 OD025315; 1 OT2 OD025337; 1 OT2 OD025276. In addition, the *All of Us* Research Program would not be possible without the partnership of its participants.

## Notes

### Competing Interest Statement

The authors have declared no competing interest.

### Author Declarations

Approval to use UK Biobank individual-level in this work was obtained under application 33297 at http://www.ukbiobank.ac.uk. Approval to use All of Us controlled tier data in this work was obtained through application at https://www.researchallofus.org.

