## Supplementary Figures and Note for "Calibrated prediction intervals for polygenic scores across diverse contexts"

(see next page)

**Figure S1: Relative $\Delta R^2$ for "white British" and all individuals in UK Biobank.** Numerical values of relative $\Delta R^2$ are displayed for trait-context pairs with statistically significant differences (multiple testing correction for all 72×11 trait-context pairs in this figure; $p < 0.05 / (72\times11)$). '*' are displayed for context-trait pairs with nominally significant differences (multiple testing correction for 11 contexts; $p < 0.05 / 11$). See Figure 3 caption for more details. Numerical results are reported at Table S2.

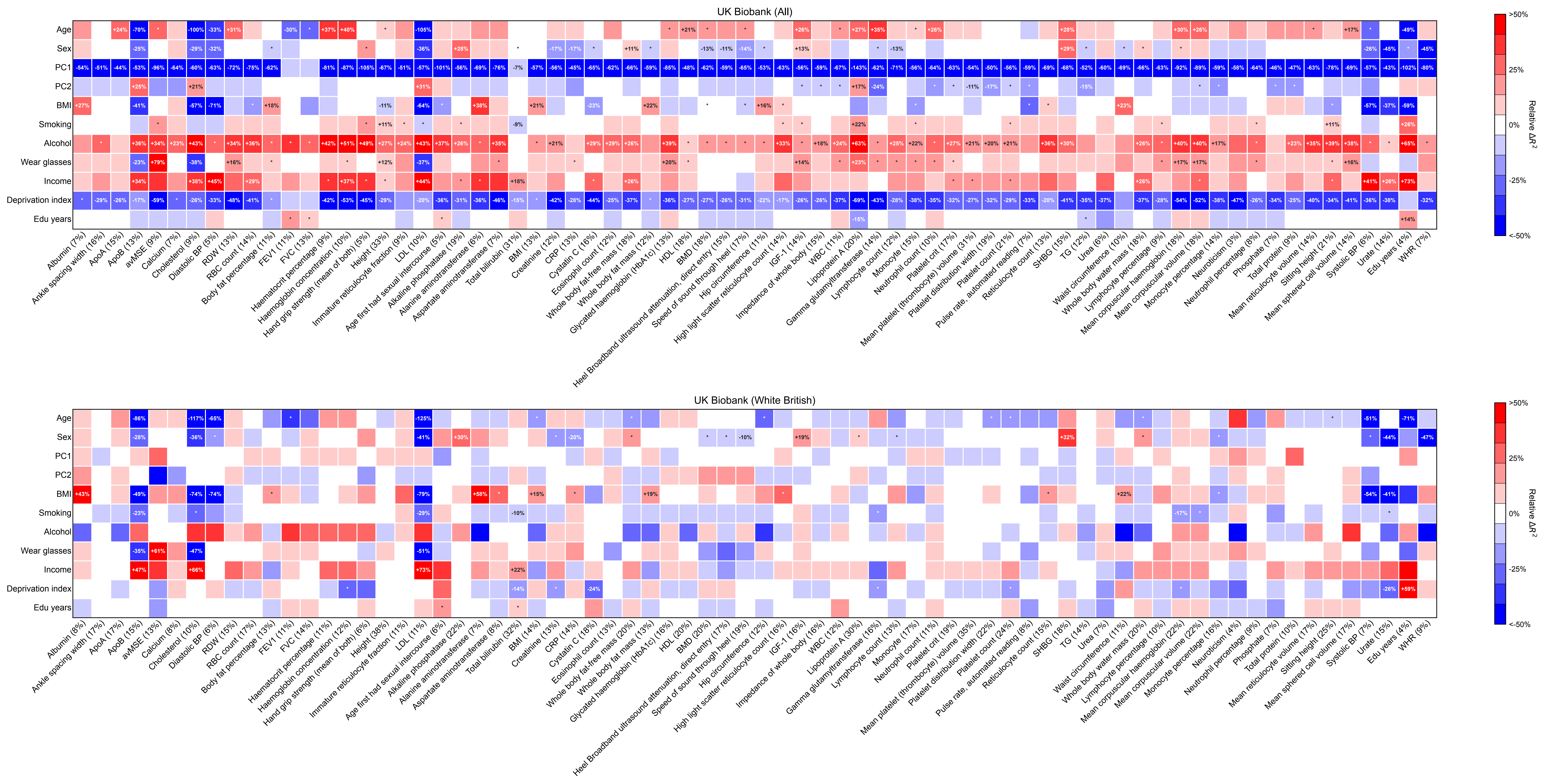
UK Biobank (All)
Age
Sex
PC1
PC2
BMI
Smoking
Alcohol
Wear glasses
Income
Deprivation index
Edu years
Relative ΔR²
>50%
25%
0%
-25%
<-50%
UK Biobank (White British)

(see next page)
**Figure S2: Estimated $\beta_\sigma$ for "white British" and all individuals in UK Biobank.** We show estimated $\beta_\sigma$ in CalPred model. Numerical results are reported at Table S2.

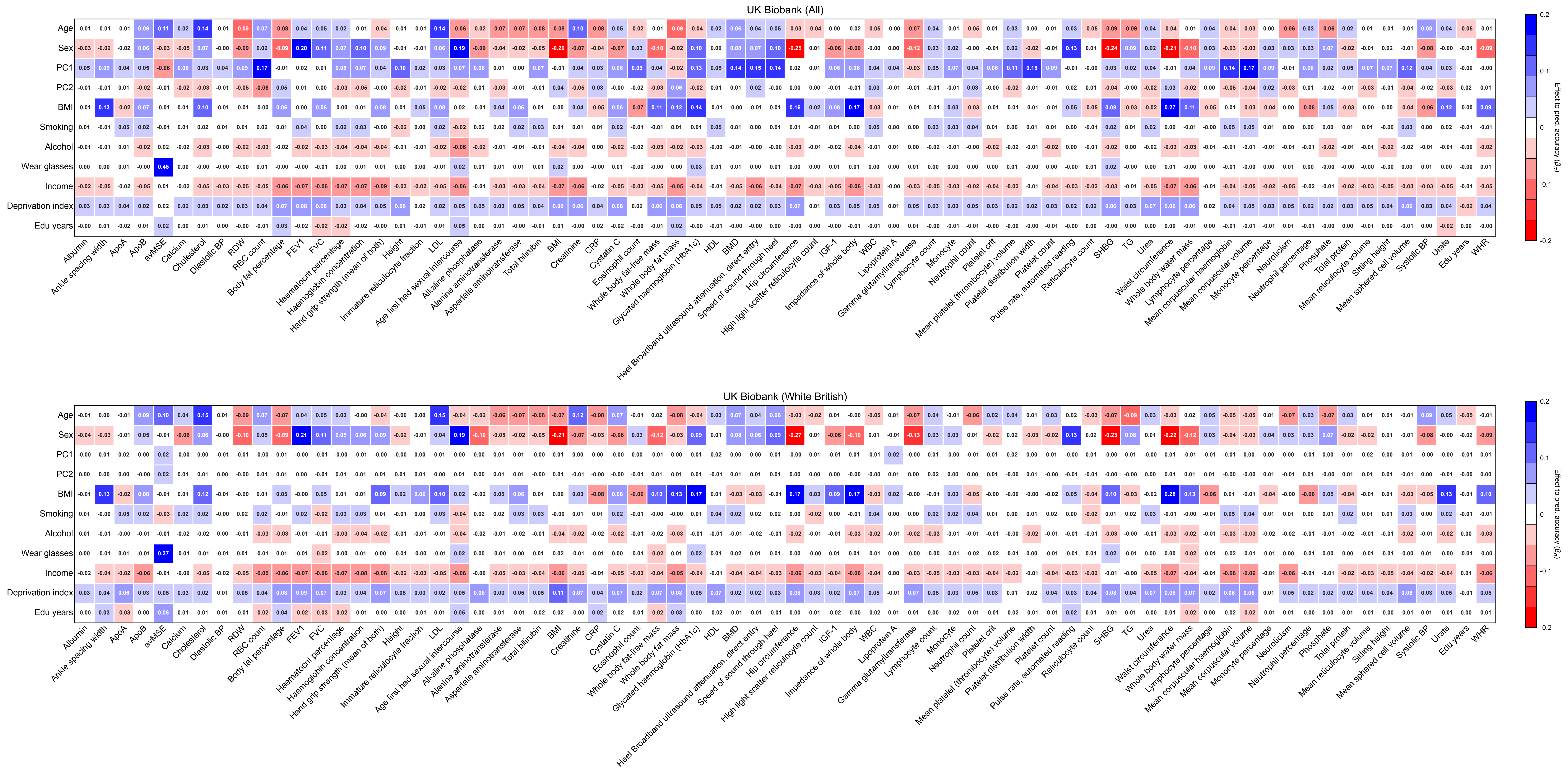
UK Biobank (All)
UK Biobank (White British)
Age
Sex
PC1
PC2
BMI
Smoking
Alcohol
Wear glasses
Income
Deprivation index
Edu years
Albumin
Ankle spacing width
ApoA
ApoB
avMSE
Calcium
Cholesterol
Diastolic BP
RDW
RBC count
Body fat percentage
FEV1
FVC
Haematocrit percentage
Haemoglobin concentration
Hand grip strength (mean of both)
Height
Immature reticulocyte fraction
Age first had sexual intercourse
LDL
Alkaline phosphatase
Alanine aminotransferase
Aspartate aminotransferase
Total bilirubin
BMI
Creatinine
CRP
Cystatin C
Eosinophil count
Whole body fat-free mass
Whole body fat mass
Glycated haemoglobin (HbA1c)
HDL
BMD
Heel Broadband ultrasound attenuation, direct entry
Speed of sound through heel
Hip circumference
High light scatter reticulocyte count
IGF-1
Impedance of whole body
WBC
Lipoprotein A
Gamma glutamyltransferase
Lymphocyte count
Monocyte
Neutrophil count
Platelet crit
Mean platelet (thrombocyte) volume
Platelet distribution width
Platelet count
Pulse rate, automated reading
Reticulocyte count
SHBG
TG
Urea
Waist circumference
Whole body water mass
Lymphocyte percentage
Mean corpuscular haemoglobin
Mean corpuscular volume
Monocyte percentage
Neuroticism
Neutrophil percentage
Phosphate
Total protein
Mean reticulocyte volume
Sitting height
Mean sphered cell volume
Systolic BP
Urate
Edu years
WHR
Effect to pred. accuracy ($\beta_s$)
0.2
0.1
0
-0.1
-0.2

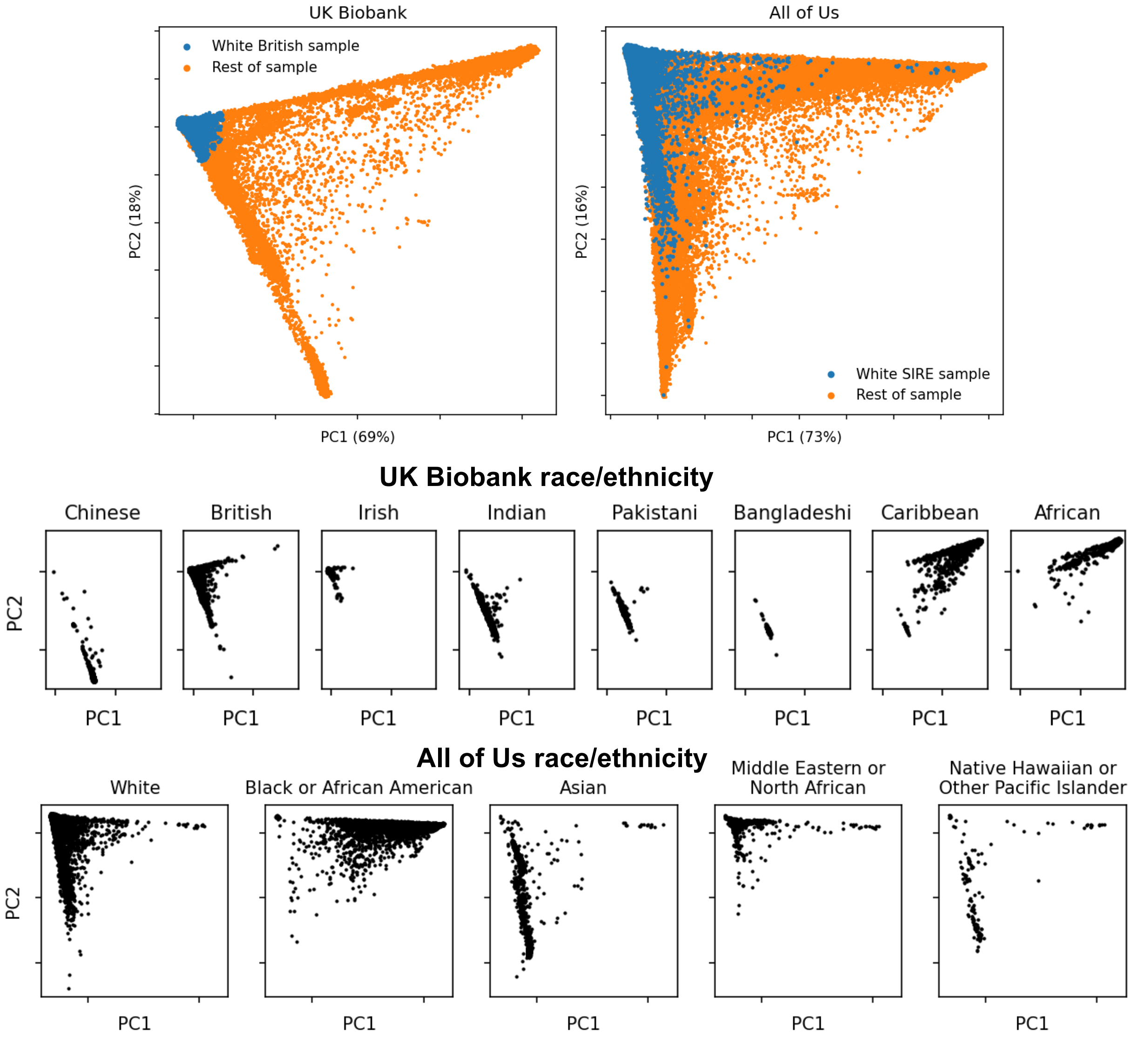


**Figure S3: Principal components calculated in UK Biobank and All of Us.** PC1 and PC2 were calculated in all individuals in UK Biobank and in All of Us. We show self-reported race/ethnicity in UK Biobank and All of Us to help interpret PC1/PC2. We show the proportion of variance explained by PC1 and PC2 out of top ten PCs calculated in each dataset in parentheses.

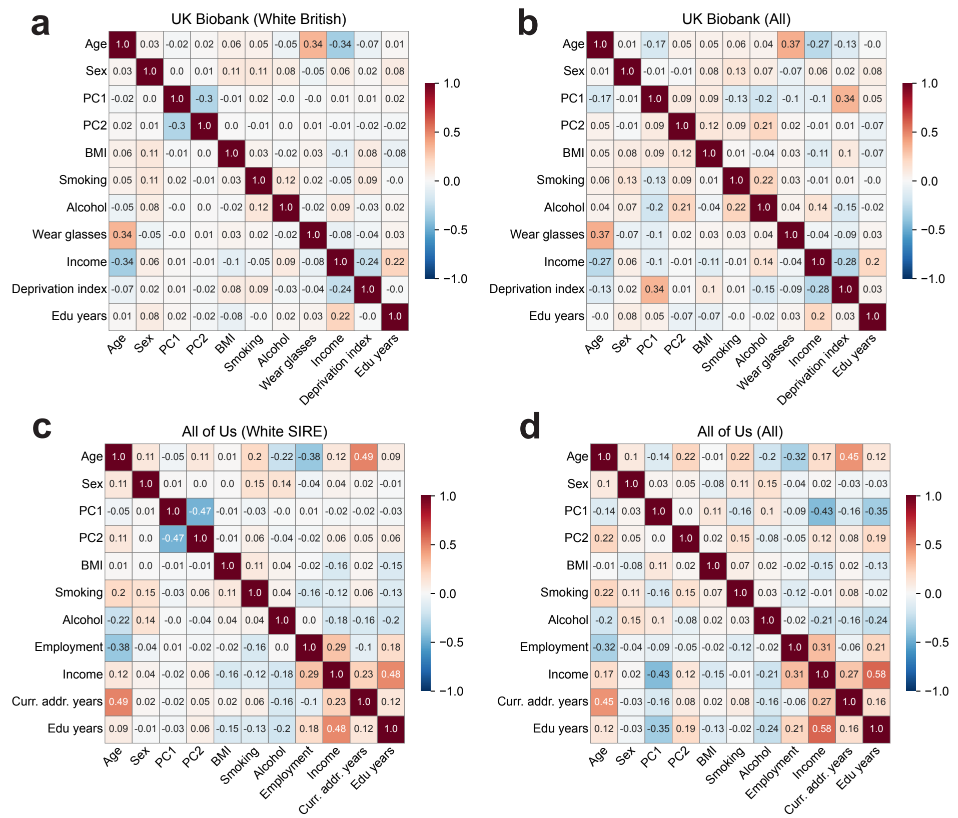


**Figure S4: Pearson's correlation between covariates in UK Biobank and All of Us datasets.** Pearson's correlations were calculated separately within individuals annotated with "white British" in UK Biobank and within individuals with SIRE "white" in All of Us **(a,c)** and across all individuals **(b,d)**

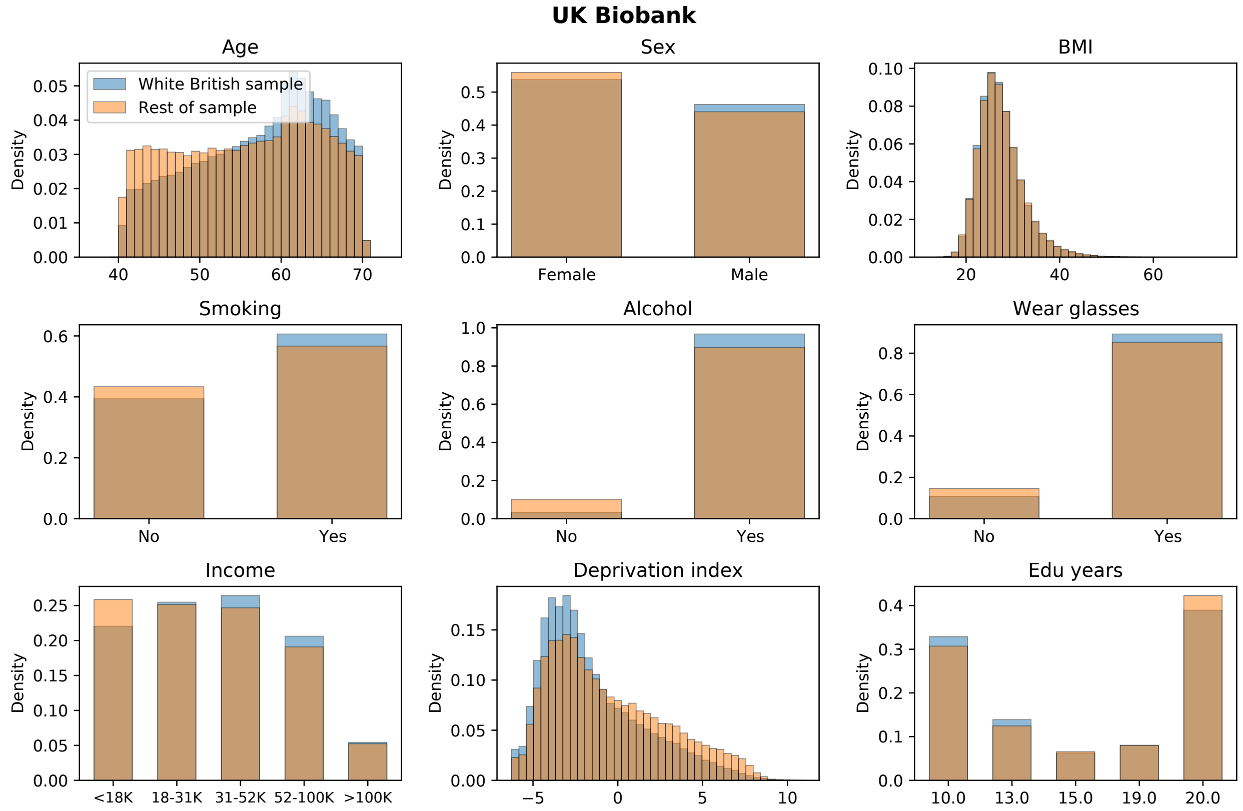


**Figure S5. Distribution of environmental context variables in UK Biobank.** We show context distribution separately for "white British" individuals and rest of individuals in UK Biobank.

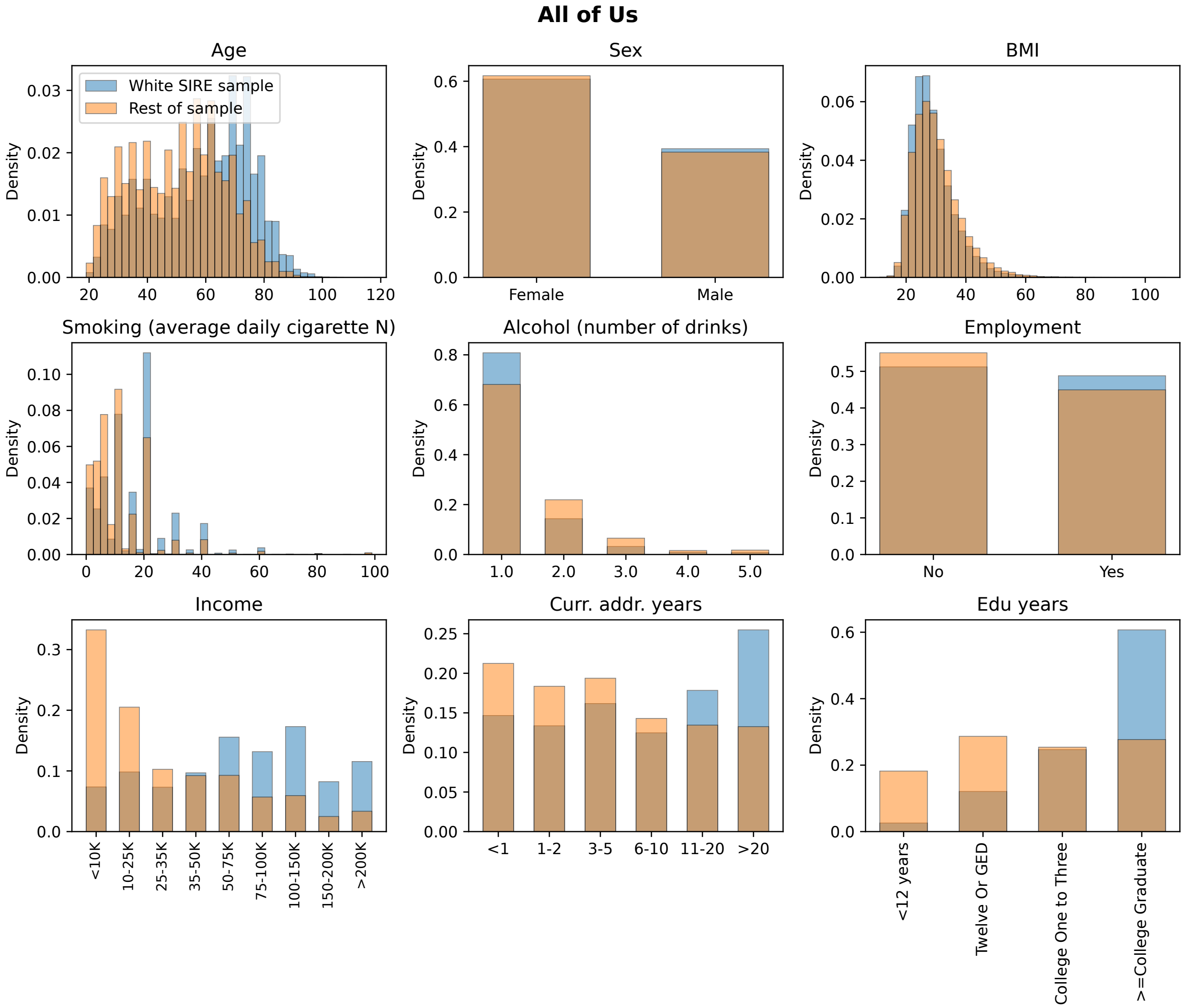


**Figure S6. Distribution of environmental context variables in All of Us.** We show context distribution separately for "white SIRE" individuals and other individuals in All of Us.

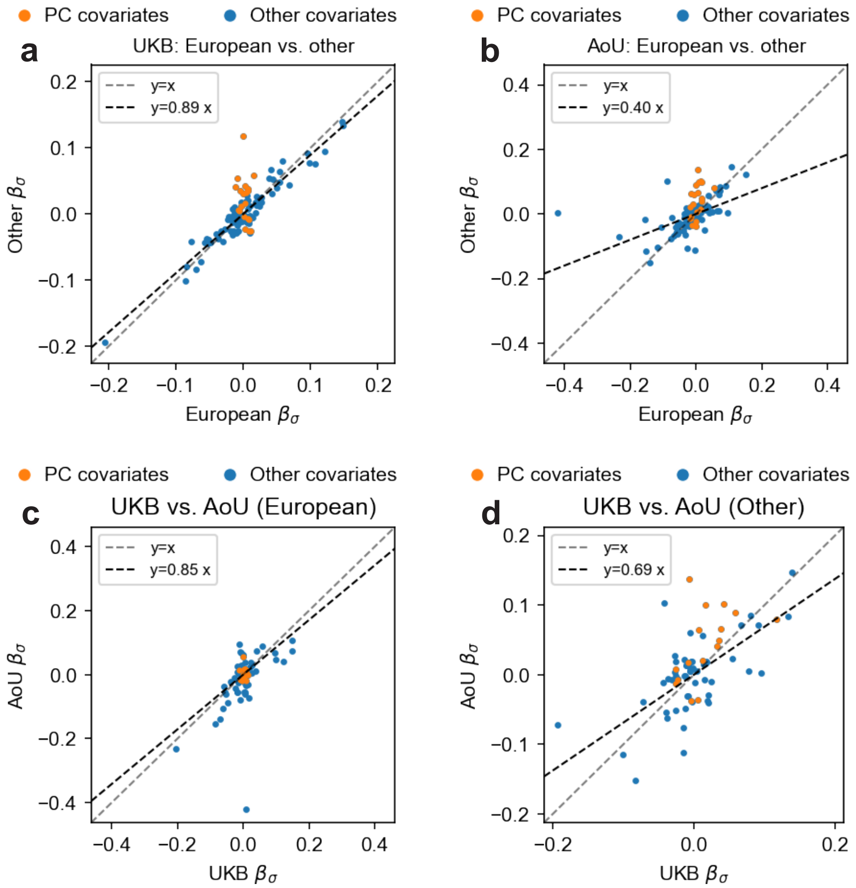


**Figure S7: Comparison of fitted parameters across populations and biobanks.** We compare the estimated $\beta_\sigma$ across populations **(a-b)** and biobanks **(c-d)**. Each dot denotes a trait-context pair. We separately annotate genetic ancestry contexts (using principal components) and other contexts, because PC contexts only have small variations within Europeans ("white British" in UK Biobank or "white SIRE" in All of Us), therefore are not expected to be comparable between European and other populations. The regression slope in each figure is calculated across all estimated $\beta_\sigma$. For **(c-d)**, we include traits that are shared across biobanks. Overall, we find that the fitted parameters are highly consistent across populations and biobanks.

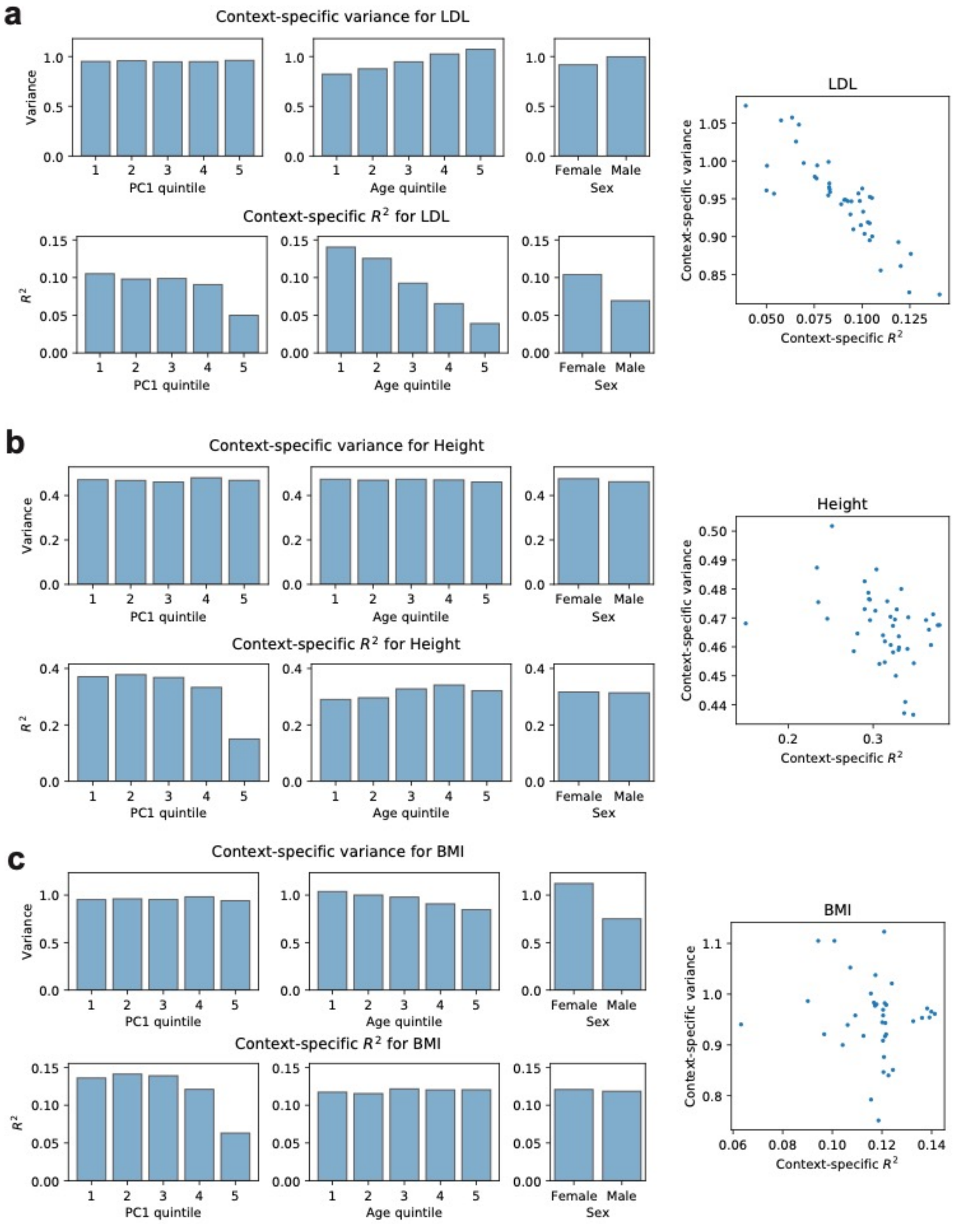


**Figure S8. $R^2$ and phenotypical variance in context strata for example traits.** We calculate $R^2$ and phenotypical variance by PC1 quintile, age quintile and sex for three example traits of LDL, height and BMI in all individuals from UK Biobank**.** We determined that variable $R^2$ across contexts were not solely driven by differences of phenotype variance in context strata. The relationship between $R^2$ and phenotypical variance depends on the specific trait-context being studied. For example, height $R^2$ varies across age quintiles while the phenotypical variance remains relatively constant; BMI $R^2$ stays relatively constant across age quintiles while the phenotypical variance varies.

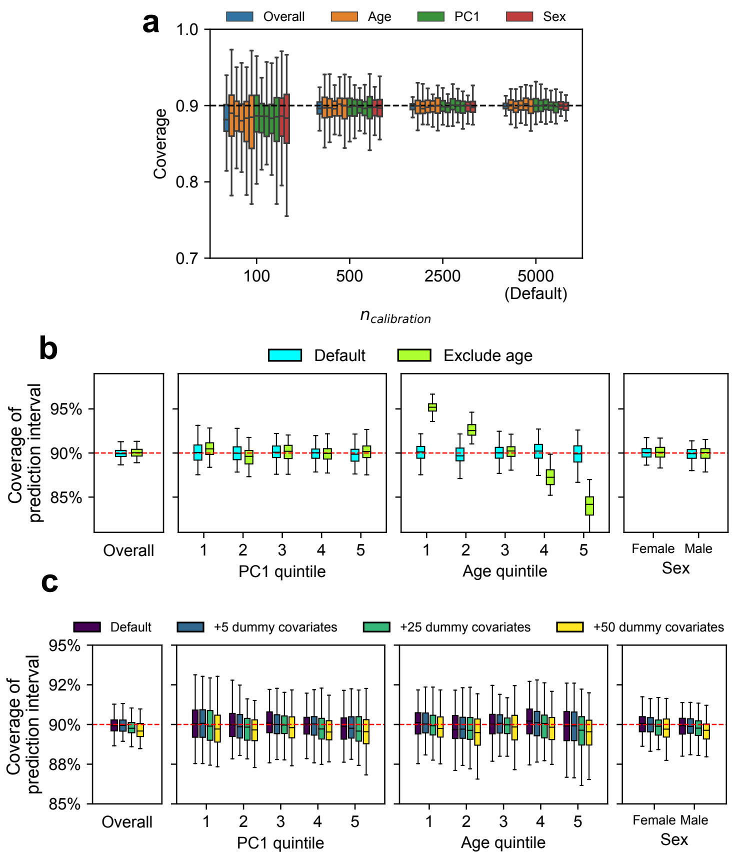


**Figure S9: Additional simulation results (related to Figure 5).** We performed simulations to investigate factors that influence coverage of prediction intervals. We compared coverage in these alternative scenarios with default scenario (marked by 'Default' in the figure) where we performed calibration using age, PC1, and sex and 5000 individuals as calibration data (same as Figure 5). **(a)** Coverage of prediction intervals with varying number of individuals used in calibration ($N_{cal}$ = 100, 500, 2500, 5000). We evaluated the coverage both at the overall level and within each group (groups are denoted by colors) using 5,000 testing individuals. Different box plots with the same color denotes different strata for each context (quintile for age and PC1; male/female for sex). We determined coverages had more downward bias and higher variance when less individuals are used in the calibration. **(b)** Coverage of prediction intervals when certain covariates (contexts) were unmeasured. To simulate unmeasured covariate, we performed calibration using PC1 and sex only (excluding age). And we determined prediction intervals were mis-calibrated along the unmeasured context of age in this scenario. **(c)** Coverage of prediction intervals when including excessive dummy covariates when performing calibration. We simulated dummy variables with no effects to phenotype variance (number of dummy covariates $N_{dummy}$ = 5, 25, 50; drawn from N(0,1)) and included them in calibration to investigate the effect of including excessive covariates to the coverage. We determined coverages had more downward bias and higher variance when more dummy variables were used in the calibration.

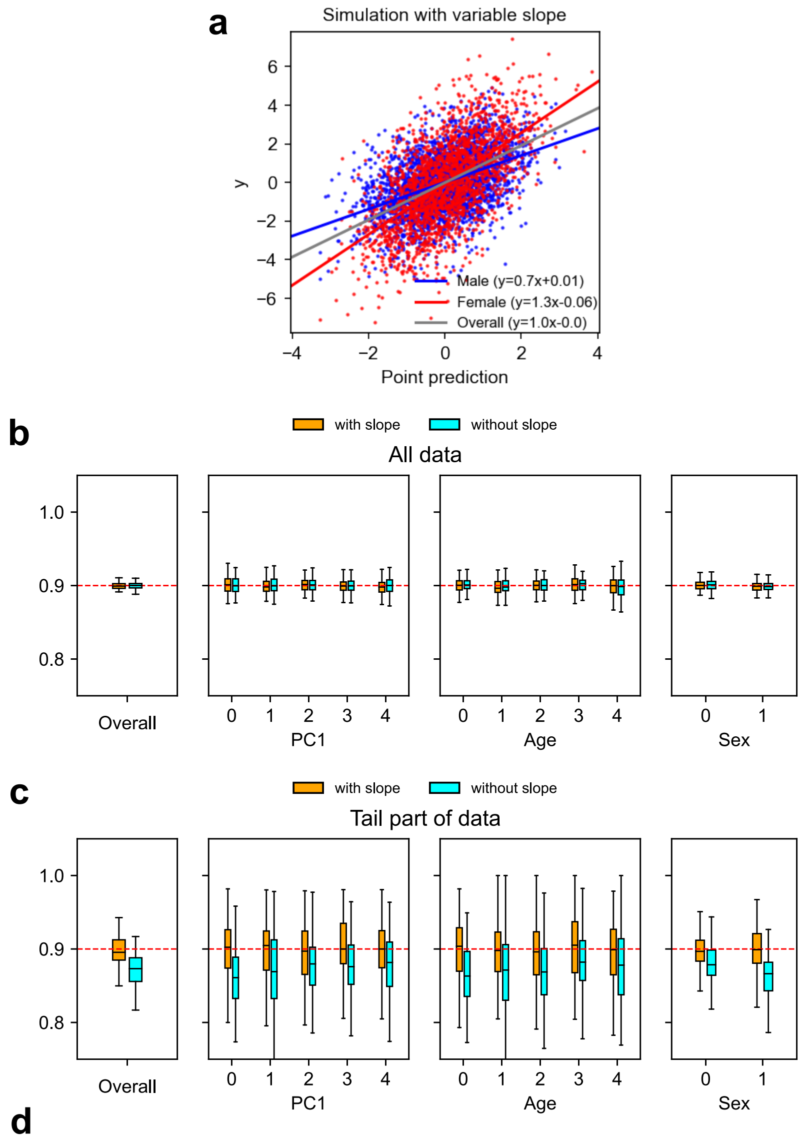


**d**

| | Constant slope | | Variable slope (with slope) | | Variable slope (without slope) | |
|---|---|---|---|---|---|---|
| | True | 95% CI | True | 95% CI | True | 95% CI |
| Age var | 0.25 | [0.244, 0.252] | 0.25 | [0.244, 0.252] | 0.25 | [0.234, 0.243] |
| Sex var | 0.20 | [0.192, 0.200] | 0.20 | [0.192, 0.200] | 0.20 | [0.209, 0.217] |
| PC1 var | 0.15 | [0.143, 0.153] | 0.15 | [0.143, 0.153] | 0.15 | [0.139, 0.148] |
| Age slope | 0.00 | [-0.005, 0.004] | 0.00 | [-0.005, 0.004] | 0.00 | NA |
| Sex slope | 0.00 | [-0.005, 0.004] | 0.30 | [0.296, 0.305] | 0.30 | NA |
| PC1 slope | 0.00 | [-0.006, 0.003] | -0.05 | [-0.056, -0.047] | -0.05 | NA |
| Prediction beta | 1.00 | [0.994, 1.003] | 1.00 | [0.994, 1.003] | 1.00 | [0.937, 0.946] |

**Figure S10: Simulations with varying slopes (between phenotypes and point predictions).** We simulated variable slopes (in regressing phenotypes against point predictions) in addition to the variable variances (as in Figure 5). We applied CalPred with or without fitting the slope parameters (denoted by 'with slope' / 'without slope') and evaluated the coverage of prediction intervals. We simulate point predictions as $y \sim \mathcal{N}(\hat{y} \times (1 + \sum \beta_{\eta,c} \times c), \exp(\beta_{\sigma,0} + \sum_c \beta_{\sigma,c} \times c)$, where true $\beta_{\eta,\text{age}} = 0, \beta_{\eta,\text{sex}} = 0.3, \beta_{\eta,\text{PC1}} = -0.05$. Other simulation settings were the same as in Figure 5. **(a)** Example data with variable slope. Because $\beta_{\eta,\text{sex}} = 0.3$, slopes between phenotype and point predictions were different between male and female. When the interaction parameters $\beta_\eta$ were not fitted in the calibration, an overall fitted line was obtained. Consequently, at the extreme of point predictions, prediction intervals would have lower-than-expected coverages because of the shift in the mean predictions. **(b)** Coverage of prediction intervals evaluated across all 5,000 target individuals. CalPred produced prediction intervals with expected coverage level at the overall level regardless of whether interaction term is fitted or not. **(c)** Coverage was evaluated across 5% individuals at the tail distribution of point predictions (left tail~2.5%, right tail ~2.5%). When applied without fitted slope, prediction intervals had lower-than-expected coverage as a result of biased point predictions. When applied with fitted slope, prediction intervals resumed well-calibration at extremes of the distribution. **(d)** Numerical results of parameter estimation. We report the true parameter values and 95% confidence intervals of estimated parameters across 100 simulations. 'Constant slope' column denotes simulations in Figure 5. 'Variable slope (with slope)' / 'Variable slope (without slope)' denotes simulations in **(a-c)** with / without fitting interaction term. We determined that when the model is correctly specified ('Constant slope' and 'Variable slope (with slope)'), parameter estimation was unbiased. When the model is mis-specified ('Variable slope (without slope)'), parameter estimation remained robust.

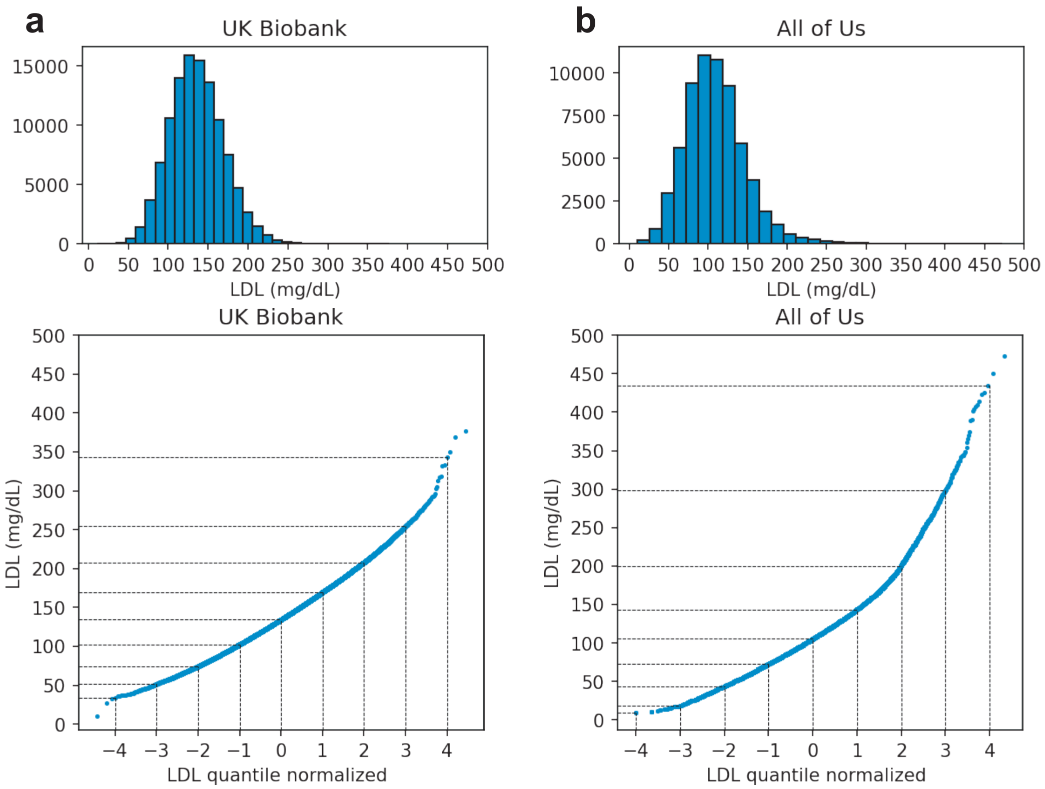


**Figure S11: correspondence between quantile normalized and raw LDL levels for UK Biobank (a) and All of Us (b).** We plot distribution of LDL in raw unit of mg/dL in upper panels. We also plot the correspondence between quantile normalized LDL (to normal distribution of zero mean and unit variance) and original LDL measurement in unit of mg/dL. We note that distribution of LDL is right-skewed in both UK Biobank and All of Us. Therefore, the same unit increase in quantile normalized scale at different LDL levels can correspond to different amount of change in the original LDL measurement.

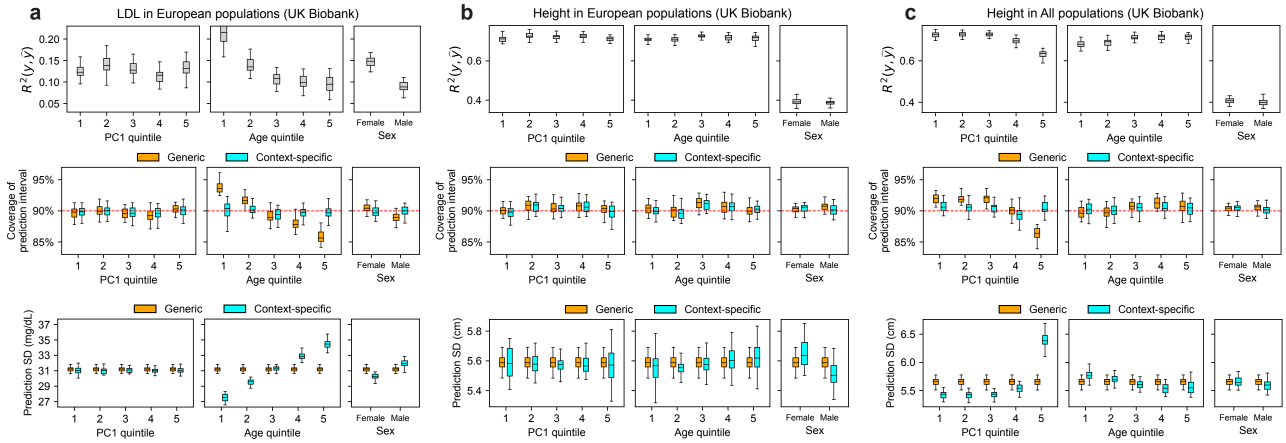


**Figure S12: Results for other populations and traits in UK Biobank (related to Figure 6).** We plot results for LDL and height in "white British", and height in all individuals. **(top panel)** prediction $R^2$ between phenotype and point predictions **(middle panel)** coverage of generic vs. context-specific 90% prediction intervals **(bottom panel)** average length of generic vs. context-specific 90% prediction intervals in each context. See Figure 6 caption for more details.

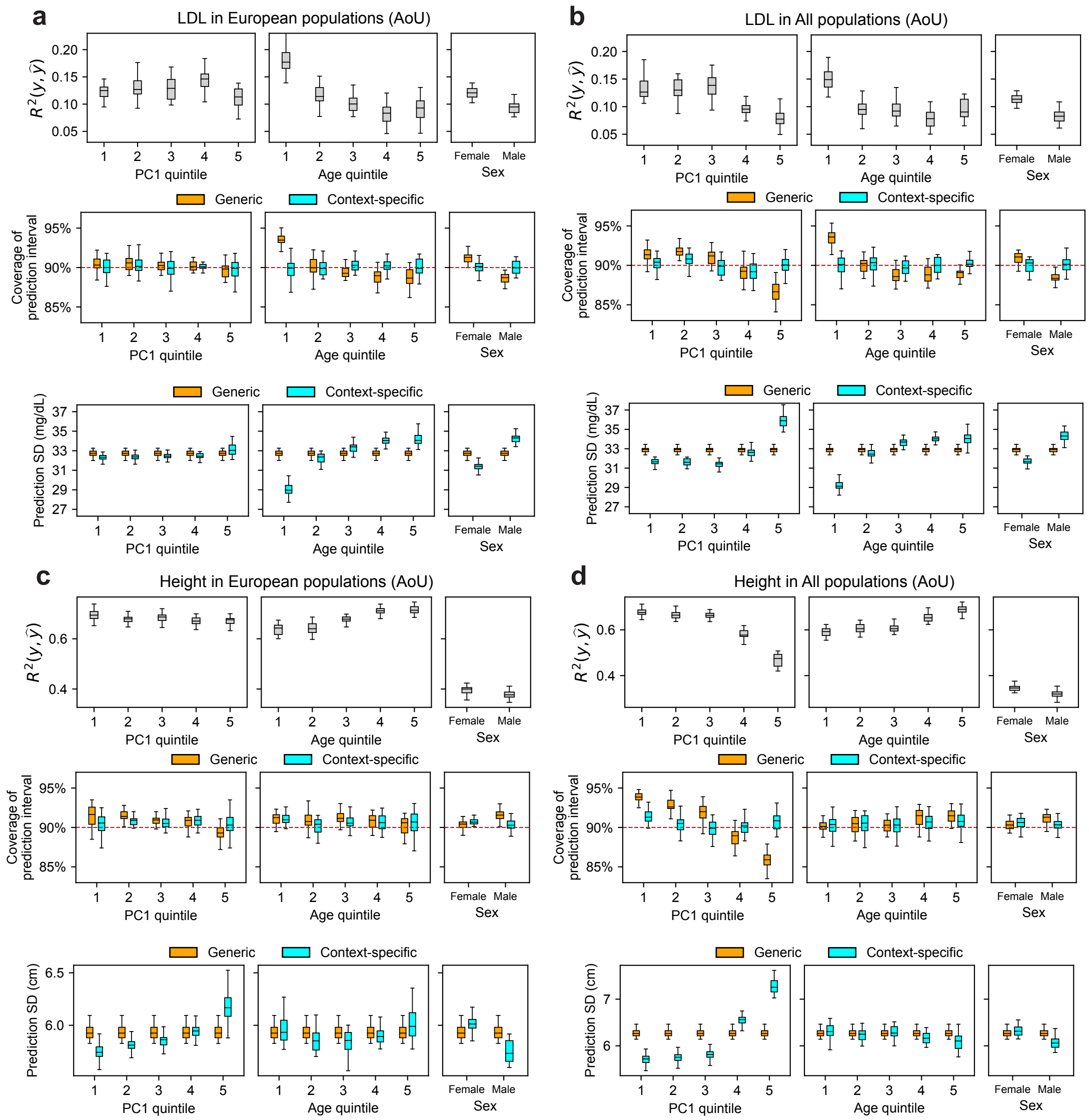


**Figure S13: Results for other traits in All of Us (related to Figure 6).** We plot results for LDL and height in "white SIRE" and height in all individuals. **(top panel)** prediction $R^2$ between phenotype and point predictions **(middle panel)** coverage of generic vs. context-specific 90% prediction intervals **(bottom panel)** average length of generic vs. context-specific 90% prediction intervals in each context. See Figure 6 caption for more details. Interestingly, while patterns of $R^2$ variation across PC1 were similar in UK Biobank and All of Us, generic intervals had more stable coverages across PC1 in UK Biobank compared to All of Us. This can be explained by that part of variable $R^2$ in UK Biobank being induced by the varying phenotype-predictor regression slope across PC1 in addition to variable noise level, which can be partially modeled through PC1 interaction terms (Supplementary Note).

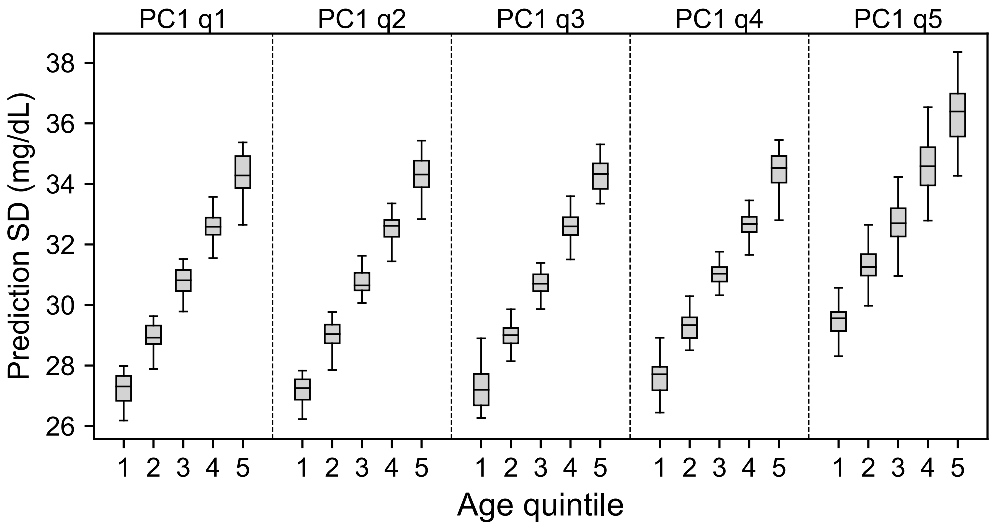


**Figure S14: Length of prediction interval in predicting LDL as a function of PC1 and age (related to Figure 6).** Average length of context-specific 90% prediction intervals stratified by both five quintiles of PC1 and five quintiles of age. By contrasting individuals with youngest age and smallest PC1 (leftmost) quintiles versus those of oldest age and largest PC1 quintiles (rightmost), we find larger differences of prediction interval length across these subgroups compared to results obtained when single context (either age or PC1) is considered as in Figure 6. By considering the contribution of both age and PC1 (largest two contributors to context-specific accuracy), we detected larger differences for individuals with youngest age and smallest PC1 quintiles (more similar to European) versus those of oldest age and largest PC1 quintiles (less similar to European) (27.3 vs. 36.3 mg/dL, 33% difference). Each box plot contains data across 30 random samples with each sample of 5,000 training individuals and 5,000 target individuals (30 points for each box plot), the center corresponds to the median; the box represents the first and third quartiles of the points; the whiskers represent the minimum and maximum points located within 1.5× interquartile range from the first and third quartiles, respectively.

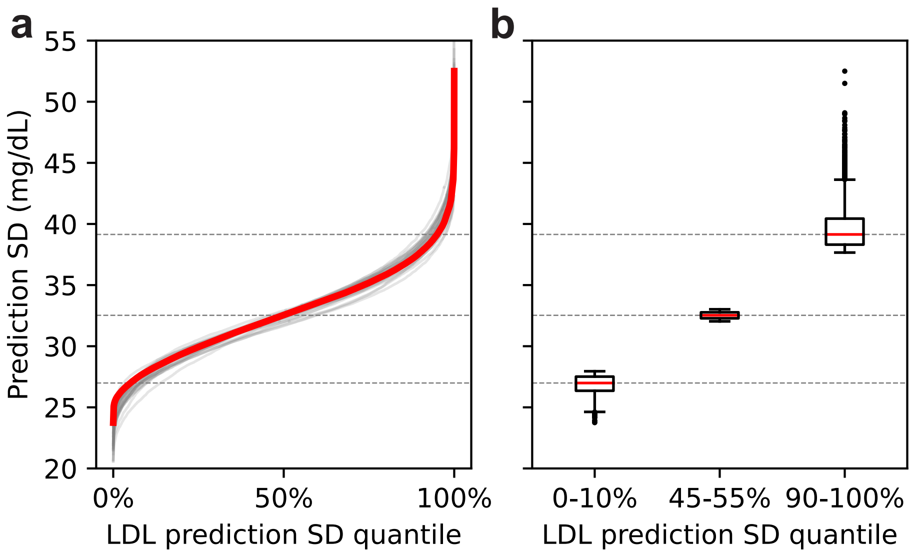


**Figure S15. Variation of prediction SD accounting for all contexts in All of Us (related to Figure 6). (a)** Ordered LDL prediction SD in unit of mg/dL. Gray lines denote prediction SD obtained with random sample of 5,000 training and applied to 5,000 testing individuals. Red line denote prediction SD obtained from all individuals. **(b)** Box plots of results in (a) from individuals of LDL prediction SD quantile of 0-10%, 45-55%, 90-100%; the center corresponds to the median; the box represents the first and third quartiles of the points; the whiskers represent the minimum and maximum points located within 1.5× interquartile range from the first and third quartiles, respectively.

### Supplementary Tables

**Table S1: Trait information in UK Biobank.** We report trait names, sample size used in training PGS weights, estimated heritability in the training sample, prediction $R^2$ (between PGS and residual phenotypes with covariates regressed out) and sample sizes used in testing populations, separately for white British and all populations. These traits are selected for their sufficient predictive power and/or biological importance.

**Table S2: Numerical results of relative $\Delta R^2$ and estimated $\boldsymbol{\beta}_\sigma$ in UK Biobank.** We report trait, context, relative $\Delta R^2$ differences between groups, and z-score for the significance of $R^2$ differences between groups. We also report $\boldsymbol{\beta}_\sigma$ estimates and corresponding SEs.

**Table S3: Trait information in All of Us.** We report trait names, prediction $R^2$ (between PGS and residual phenotypes regressed out of covariates) and sample size used in testing populations, separately for self-reported race of white and all populations.

**Table S4: Numerical results of relative $\Delta R^2$ and estimated $\boldsymbol{\beta}_\sigma$ in All of Us.** We report trait, context, relative $\Delta R^2$ differences between groups, and z-score for the significance of $R^2$ differences between groups. We also report $\boldsymbol{\beta}_\sigma$ estimates and corresponding SEs.

### Supplementary Note

**$R^2$ is impacted by both variable slope and variable noise.**
We consider a model where phenotype $y$ is a function of predicted phenotype $\hat{y}$ multiplied by a slope $s$ in additional to environmental noise $e$: $y = s \cdot \hat{y} + e$, and we discuss how $R^2(y, \hat{y})$ varies as a function of both $s$ and $\mathrm{Var}[e]$. We consider a scenario where slope can vary as a function of contexts, for example, the PGS-phenotype regression slope $s$ can vary across sex (see ref.[15]). Under such scenarios, the prediction accuracy is

$$R^2(y, \hat{y}) = R^2(s \cdot \hat{y} + e, \hat{y}) = \frac{s^2 \cdot \mathrm{Var}[\hat{y}]}{s^2 \cdot \mathrm{Var}[\hat{y}] + \mathrm{Var}[e]}.$$

Therefore, holding $\mathrm{Var}[\hat{y}]$ as constant, $R^2(y, \hat{y})$ increases with increasing $s$ and decreasing $\mathrm{Var}[e]$. In other words, variable $R^2$ can stem from variable slope $s$, even when $\mathrm{Var}[e]$ is constant (prediction interval length is therefore also constant). CalPred models $\mathrm{Var}[e]$ as a function of covariates and therefore can capture the variable $R^2$ due to change in $\mathrm{Var}[e]$. And we note that the variable $R^2$ due to variable slope $s$ can be captured by modeling variable slope in point prediction terms in CalPred model.

We analyzed LDL in UK Biobank to demonstrate these reasonings in practice. We randomly selected 5,000 individuals as training sample to train CalPred model and 5,000 individuals as target sample to perform the evaluation. Random sampling was repeated 30 times. We evaluated prediction accuracy $R^2$, standard deviation of the residuals, standard deviations of the predicted phenotype, and regression slope in regression of phenotype against predicted phenotype $\hat{y}$ following similar settings in Figure 6. Results are shown in the following tables.

| PC1 q | R2 | std(resid) | std(pred) | std(y) | slope |
|---|---|---|---|---|---|
| 1 | 0.148(0.004) | 0.913(0.003) | 0.376(0.002) | 0.989(0.004) | 1.056(0.014) |
| 2 | 0.139(0.004) | 0.922(0.005) | 0.379(0.002) | 0.994(0.005) | 1.012(0.015) |
| 3 | 0.14(0.003) | 0.914(0.004) | 0.375(0.002) | 0.986(0.004) | 1.022(0.013) |
| 4 | 0.137(0.004) | 0.924(0.004) | 0.373(0.002) | 0.995(0.004) | 1.017(0.014) |
| 5 | 0.087(0.004) | 0.952(0.003) | 0.37(0.002) | 0.997(0.003) | 0.817(0.017) |

| PC1 q | R2 | std(resid) | std(pred) | std(y) | slope |
|---|---|---|---|---|---|
| 1 | 0.151(0.004) | 0.911(0.003) | 0.383(0.003) | 0.989(0.004) | 1.026(0.015) |
| 2 | 0.14(0.004) | 0.922(0.005) | 0.385(0.003) | 0.994(0.005) | 0.978(0.014) |
| 3 | 0.142(0.003) | 0.913(0.004) | 0.382(0.003) | 0.986(0.004) | 0.992(0.013) |
| 4 | 0.138(0.004) | 0.923(0.004) | 0.377(0.002) | 0.995(0.004) | 0.993(0.014) |
| 5 | 0.085(0.004) | 0.953(0.003) | 0.331(0.004) | 0.997(0.003) | 0.894(0.021) |

**Results for predicting LDL stratified by PC1.** We show prediction accuracy $R^2(y, \hat{y})$, standard deviation of the residuals in regression ($\mathrm{std}(y - \hat{y})$), standard deviation of predicted phenotype, standard deviation of phenotype, and regression slope of $y$ against $\hat{y}$. Regression was separated performed in each subset of the data stratified by PC1. **Upper panel:** not including PGS interaction terms. **Lower panel:** including PGS interaction terms.

First, as already shown in Figure 6, prediction accuracy $R^2$ changed drastically along PC1, especially when comparing 5$^{th}$ quintile and 1$^{st}$ quintile of PC1. The standard deviation of the residuals (corresponding to $\mathrm{Var}[e]$ term and prediction interval length), however, did not change drastically along PC1. This was consistent with the observation that constant generic prediction intervals provided reasonable coverage for predicting LDL in UK Biobank. By contrasting the regression slope of $y$ against $\hat{y}$ across PC1 quintile, we determined that the varying $R^2$ of LDL prediction along PC1 was mainly due to different regression slope. For example, the effect of PGS was different across subpopulations stratified by PC1.

Next, we reason that including interaction in point prediction terms can better model varying slope. We included interaction term of PGS*age, PGS*sex, PGS*PC1, …, PGS*PC4 into point predictions. We found that the bias of slope is smaller (less deviated from one) and other metrics were not affected. Such bias from slope can be possibly further reduced by introducing additional parameters to model more complicated interactions among predictors.

Finally, we showcase another example where varying slope by context plays a role in varying prediction accuracy and prediction interval length. We consider the scenario to predict BMI in European populations from UK Biobank, and evaluated results across male and female.

| **Sex** | **R2** | **std(resid)** | **std(pred)** | **std(y)** | **slope** |
|---|---|---|---|---|---|
| **Female** | 0.144(0.002) | 0.999(0.002) | 0.369(0.003) | 1.08(0.002) | 1.095(0.012) |
| **Male** | 0.139(0.002) | 0.813(0.003) | 0.358(0.003) | 0.876(0.003) | 0.903(0.01) |

| **Sex** | **R2** | **std(resid)** | **std(pred)** | **std(y)** | **slope** |
|---|---|---|---|---|---|
| **Female** | 0.144(0.002) | 0.999(0.002) | 0.413(0.003) | 1.08(0.002) | 0.979(0.012) |
| **Male** | 0.139(0.002) | 0.813(0.003) | 0.326(0.003) | 0.876(0.003) | 0.993(0.012) |

We observe that the prediction accuracy $R^2$ were similar for female versus male. However, we note that their residual noise levels were drastically different (corresponding to $\mathrm{Var}[e]$ term and different prediction interval length for male and female). The reason for the absence of differential prediction accuracy was that the varying residual noise was compensated by the varying slope due to interaction of PGS effect with sex. In other words, although both slope $s$ and $\mathrm{Var}[e]$ in $\frac{s^2 \cdot \mathrm{Var}[\hat{y}]}{s^2 \cdot \mathrm{Var}[\hat{y}] + \mathrm{Var}[e]}$ were variable, $R^2(y, \hat{y}) = \frac{s^2 \cdot \mathrm{Var}[\hat{y}]}{s^2 \cdot \mathrm{Var}[\hat{y}] + \mathrm{Var}[e]}$ reflecting the relative change between the two remained constant.

In conclusion, variable $R^2$ can derive from both variable slope $s$ and variable noise $\mathrm{Var}[e]$. Such variable slope $s$ can be partially addressed by including PGS*context terms. We also note that empirically, such variable slope impact little on the prediction interval length. While modeling interactions among predictors to construct better point predictions is important, we leave extensive investigation of this topic to future work.

**Investigating source of variable accuracy due to bias versus conditional variance**

With $y_i = \mathcal{N}\big(\mu(\mathbf{c}_i), \sigma^2(\mathbf{c}_i)\big)$, CalPred models the variable conditional variance through $\sigma^2(\mathbf{c}_i)$. Therefore, CalPred performance relies on the unbiasedness of prediction mean $\mu(\mathbf{c}_i)$ to properly model the conditional variance term. We investigated the bias of prediction mean by comparing distribution of phenotype values versus prediction mean across contexts. We performed the comparison for three example traits of height, BMI and LDL across all individuals in UK Biobank.

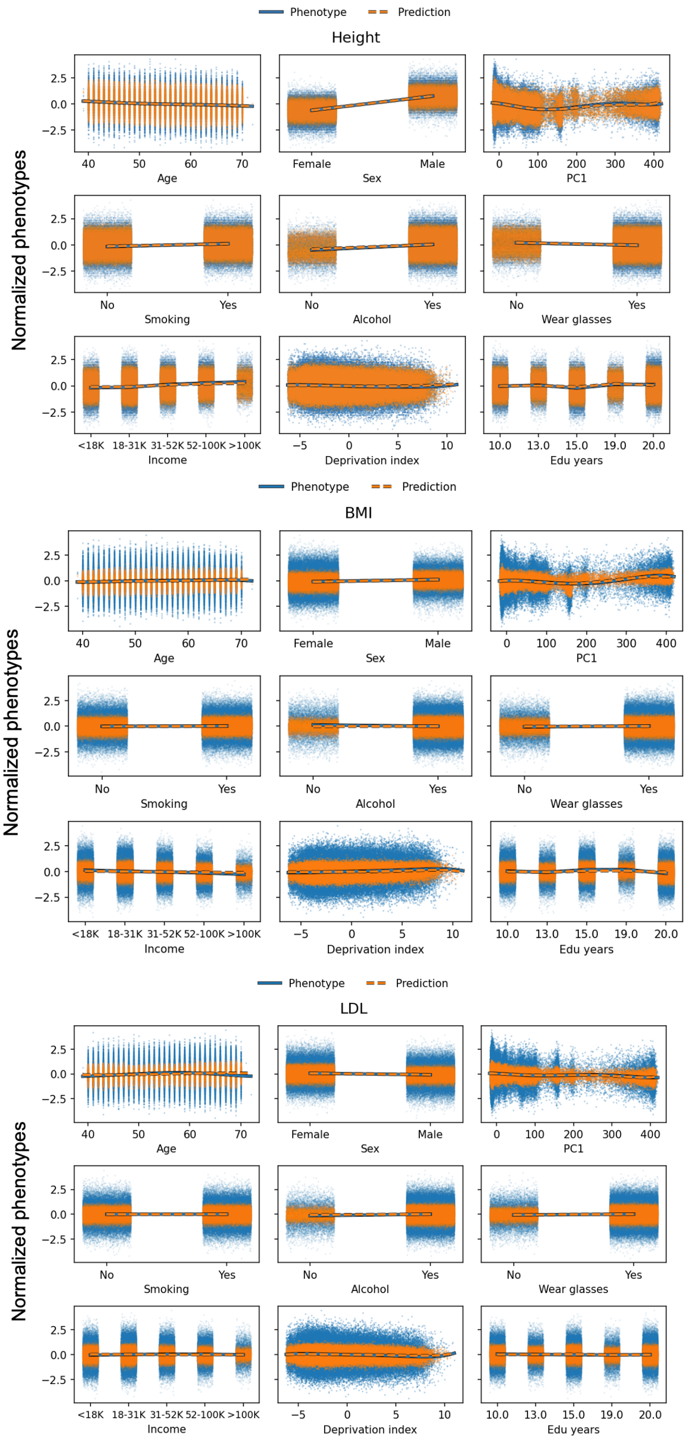


Across three example traits, we observed that the prediction mean tracked well with the phenotypical mean, indicating that prediction biases were small across context groups and that CalPred model assumption was largely valid. Meanwhile, more fine-grained modeling of prediction factors that capture more variation, for example, by modeling interactions between prediction factors will benefit CalPred model via more precise, and shorter, prediction intervals.
